# Predicting appropriateness of antibiotic treatment among ICU patients with hospital acquired infection

**DOI:** 10.1101/2023.08.15.23294109

**Authors:** Ella Rannon, Ella Goldschmidt, Daniel Bernstein, Asaf Wasserman, Dan Coster, Ron Shamir

## Abstract

Antimicrobial resistance is a growing threat to global health, leading to ineffective treatment of infection and increasing treatment failure, mortality, and healthcare costs. Inappropriate antibiotic therapy is often administered in the Intensive Care Unit (ICU) due to the urgency of treatment, but can lead to poor patient outcomes. In this study, we developed a machine learning model that predicts the appropriateness of antibiotic treatments for ICU inpatients with ICU-acquired blood infection. We analyzed data from electronic medical records (EMRs), including demographics, administered drugs, previous microbiological cultures, invasive procedures, lab measurements and vital signs. Since EMRs have high rates of missing values and since our cohort is relatively small and imbalanced, we introduced novel computational methods to address these issues. The final model achieved an AUROC of 82.8% and an AUPR of 60.6% on the training set and an AUROC score of 77.3% and an AUPR score of 40.4% on the validation set. Our study shows the potential of machine learning models for inappropriate antibiotic treatment prediction.

## Introduction

Infectious diseases are considered one of the major health risks worldwide^1^. Although the development of antimicrobial drugs has transformed the treatment of bacterial infections, the massive increase in antibiotic consumption has led to the emergence of bacterial resistance, thus reducing antibiotics efficacy^2–4^. Consequently, both the Centers for Disease Control and Prevention and World Health Organization declared antibiotic resistance as a threat to human health^5,6^, and have created guidelines for appropriate antibiotic administration^3,7,8^.

Nowadays, culture incubation is the golden standard for bacterial pathogen assessment. The process takes 48 to 72 hours. Typically, a gram stain is completed after 24 hours, organism identification is obtained after another 24 to 48 hours, and antimicrobial susceptibility testing (AST) profile is received after 72 hours^9,10^. However, since early antibiotic intervention is a critical determinant of patients’ survival, patients are often treated with empiric antibiotic therapy, where antibiotics are administered prior to the receipt of blood culture and AST results. This treatment is based on the clinician’s preliminary evaluation of the patient’s health state, infection history, and local bacterial resistance patterns^11,12^. Nevertheless, such treatment might be inappropriate, as the antibiotic administered might not be suitable to the pathogen. In particular, ICU-acquired infections are more likely to be resistant to a broad spectrum of antibiotics^13^.

Recently, it has been shown that inappropriate antibiotic therapy (IAT) is associated with higher incidence of treatment failure, higher mortality rate, and a prolonged hospital stay, which can also result in higher healthcare cost^14,15^. Moreover, in severe cases of bloodstream infections and in cases of septic shock, IAT was found to be the most important factor in ICU patients’ outcome^16^. Therefore, it is essential to develop methods for rapid identification of treatment appropriateness in ICU patients. However, to the best of our knowledge, no machine learning model has been developed for prediction of IAT in ICU patients with hospital acquired infection.

In this study, we developed a machine-learning model that predicts the appropriateness of antibiotic empirical treatments based on electronic medical records (EMRs) of ICU patients with hospital acquired infection. Our prediction is made 24 hours after the blood culture was taken and thus approximately 24 hours after the empiric antibiotic has already been administered^17^. Unlike previous models that tried to make the prediction at the time of culture collection, we assume that at the 24h point the patient’s measurements such as lab measurements and vital signs are already affected by the antibiotic intervention and can give indication whether the antibiotic treatment was appropriate.

In the process of method development, we also devised novel computational methods and a flexible pipeline to deal with challenges that often arise when dealing with EMR data, such as missing values and imbalanced data. The methods are described in detail and can be adopted for other models that use EMRs.

## Results

### Cohort Description

We used MIMIC-III, an open-access, anonymized database of EMRs of ICU patients, to develop, validate, and test our model. Data from 53,423 distinct ICU stays of adult patients admitted to Beth Israel Deaconess Medical Center (Boston, MA, USA) between 2001 and 2012 are included in the database.^18^ The dataset contains for each patient stay time-independent (static) features, such as age, gender, ethnicity, weight, height, and a large variety of time-dependent (dynamic) features that are measured during hospitalization, including vital signs, lab measurements, and drug administrations. We used 55 continuous features (**Table 1**), 7 drug features (**Supplementary Table 1**) that were created by aggregating 242 drugs into 11 drug categories (**Supplementary Table 2**), and 39 categorical features (**Supplementary Table 3**). For all features, only values recorded before the *prediction time* (henceforth abbreviated as PT), set to 24 hours after the time the blood culture was taken (abbreviated as BCT), were considered. We considered for our cohort only patients with suspected hospital acquired infection (**Figure 1A-B**). Overall, the training set consisted of 105 patients divided into two classes. The *inappropriate treatment group*, defined as those who received antibiotic treatment to which the pathogen was resistant, consisted of 22 patients. The remaining 83 patients received antibiotic treatment to which the pathogen was sensitive and therefore were included in the *appropriate treatment group*.

**Table 1.** Statistics of the continuous features used in our pipeline. For each feature, the table shows, in each class, the number of patients with the feature, the mean and standard deviation of the feature’s value, the mean and standard deviation of the duration (in hours) between measurement time and blood culture time (BCT), and the p-value of t-test results between feature values of the two classes, after FDR correction. For each feature, only the last values before prediction time were taken into account for this table. All lab measurements are blood based except the vital signs and measurements marked with U, which are urine based.

|  | Inappropriate |  |  | Appropriate |  |  |  |
| --- | --- | --- | --- | --- | --- | --- | --- |
| Feature (Unit) | N | Mean ± SD | Time from BCT | N | Mean ± SD | Time from BCT (mean ± SD) | P-value |
| Age (Years) | 22 | 69.45 ± 15.19 | | 83 | 65.52 ± 16.76 | | 0.67 |
| Admission to prediction (hours) | 22 | 186.88 ± 110.86 | | 83 | 134.39 ± 109.6 | | 0.37 |
| ICU Admission to prediction (hours) | 22 | 122.38 ± 88.24 | | 83 | 106.79 ± 97.27 | | 0.81 |
| Alanine Aminotransferase (IU/L) | 16 | 171.06 ± 283.72 | 20.11 ± 52.06 | 61 | 112.69 ± 404.41 | 31.49 ± 51.16 | 0.83 |
| Alkaline Phosphatase (IU/L) | 16 | 109.94 ± 40.41 | 27.66 ± 56.25 | 59 | 83.41 ± 34.32 | 42.4 ± 81.71 | 0.29 |
| Anion Gap (mEq/L) | 22 | 14.09 ± 3.96 | -14.65 ± 8.28 | 83 | 13.25 ± 3.57 | -11.97 ± 9.2 | 0.73 |
| Arterial pH (pH) | 22 | 7.38 ± 0.1 | -9.79 ± 27.4 | 76 | 7.42 ± 0.06 | -3.98 ± 36.56 | 0.47 |
| Aspartate Aminotransferase (IU/L) | 16 | 201.31 ± 421.97 | 20.11 ± 52.06 | 61 | 68.59 ± 100.96 | 31.49 ± 51.16 | 0.6 |
| BUN (mg/dL) | 21 | 35.52 ± 25.81 | -12.63 ± 16.87 | 82 | 33.06 ± 20.12 | -11.83 ± 9.13 | 0.91 |
| Base Excess (mEq/L) | 20 | -1.75 ± 5.28 | -13.75 ± 16.19 | 75 | 1.4 ± 4.36 | -1.68 ± 40.75 | 0.27 |
| Basophils (%) | 19 | 0.07 ± 0.15 | 86.26 ± 91.59 | 56 | 0.15 ± 0.18 | 51.71 ± 84.07 | 0.41 |
| Bicarbonate (mEq/L) | 22 | 22.23 ± 5.09 | -12.02 ± 20.4 | 83 | 25.41 ± 4.47 | -9.44 ± 17.72 | 0.24 |
| CO2 (mEq/L) | 21 | 22.52 ± 4.99 | -14.98 ± 12.54 | 81 | 26.38 ± 4.78 | -8.74 ± 32.1 | 0.24 |
| Calcium (mg/dL) | 22 | 7.9 ± 0.53 | -12.43 ± 16.41 | 82 | 8.24 ± 0.68 | -8.6 ± 19.39 | 0.27 |
| Chloride (mEq/L) | 22 | 105.68 ± 6.18 | -17.28 ± 6.79 | 83 | 103.63 ± 5.38 | -13.03 ± 7.59 | 0.51 |
| Creatine Kinase (CK) (IU/L) | 15 | 305.27 ± 639.25 | 80.76 ± 74.79 | 59 | 405.9 ± 580.84 | 61.07 ± 103.45 | 0.86 |
| Creatinine (mg/dL) | 22 | 1.47 ± 1.15 | -16.45 ± 6.65 | 81 | 1.47 ± 1.26 | -10.65 ± 10.02 | 1 |
| Eosinophils (%) | 19 | 0.49 ± 0.62 | 86.23 ± 91.6 | 59 | 0.7 ± 0.87 | 50.46 ± 83.27 | 0.63 |
| Glucose (mg/dL) | 22 | 135.23 ± 43.56 | -19.58 ± 5.63 | 83 | 143.88 ± 40.3 | -18.94 ± 5.74 | 0.77 |
| Heart Rate (BPM) | 22 | 92.36 ± 17.91 | -20.55 ± 7.17 | 83 | 91.01 ± 17.14 | -22.39 ± 3.83 | 0.93 |
| Hematocrit (%) | 22 | 29.96 ± 4.53 | -16.55 ± 6.3 | 83 | 29.83 ± 4.25 | -12.7 ± 8.43 | 0.97 |
| Hemoglobin (g/dL) | 22 | 10.1 ± 1.57 | -15.47 ± 6.22 | 83 | 10.11 ± 1.43 | -10.95 ± 8.47 | 0.99 |
| INR | 22 | 1.6 ± 0.61 | -8.0 ± 25.15 | 82 | 1.37 ± 0.44 | 7.28 ± 34.87 | 0.45 |
| Ionized Calcium (mmol/L) | 17 | 1.12 ± 0.07 | -5.38 ± 29.32 | 63 | 1.14 ± 0.08 | 0.46 ± 30.24 | 0.81 |
| Lactate (mmol/L) | 19 | 2.33 ± 1.77 | -1.77 ± 28.68 | 71 | 1.85 ± 1.19 | 20.16 ± 49.91 | 0.65 |
| Lymphocytes (B) (%) | 19 | 7.85 ± 6.25 | 86.23 ± 91.6 | 58 | 8.45 ± 6.11 | 54.39 ± 84.95 | 0.91 |
| MCH (pg) | 22 | 30.03 ± 1.78 | -11.68 ± 13.07 | 81 | 30.88 ± 1.9 | -10.55 ± 9.22 | 0.37 |
| MCHC (%) | 22 | 33.61 ± 1.74 | -15.21 ± 6.1 | 83 | 33.82 ± 1.43 | -10.65 ± 8.29 | 0.87 |
| MCV (fL) | 22 | 88.98 ± 5.07 | -15.21 ± 6.1 | 80 | 91.17 ± 4.94 | -11.08 ± 8.3 | 0.41 |
| Magnesium (mg/dL) | 22 | 2.07 ± 0.37 | -13.79 ± 16.02 | 83 | 2.04 ± 0.27 | -10.88 ± 13.66 | 0.91 |
| Monocytes (B) (%) | 19 | 3.89 ± 1.92 | 87.2 ± 90.53 | 56 | 3.86 ± 2.59 | 53.43 ± 86.25 | 0.98 |
| NBP Diastolic (mmHg) | 22 | 56.68 ± 16.74 | -20.56 ± 7.18 | 83 | 58.54 ± 13.51 | -22.46 ± 3.88 | 0.88 |
| NBP Mean (mmHg) | 22 | 78.38 ± 20.88 | -20.55 ± 7.19 | 83 | 77.64 ± 15.12 | -22.45 ± 3.84 | 0.97 |
| NBP Systolic (mmHg) | 22 | 124.55 ± 30.92 | -8.24 ± 39.91 | 83 | 121.47 ± 22.44 | -17.8 ± 18.22 | 0.9 |
| Neutrophils (%) | 19 | 80.64 ± 12.2 | 94.64 ± 88.86 | 59 | 83.02 ± 7.87 | 53.58 ± 84.73 | 0.79 |
| Oxygen Saturation (%) | 22 | 97.05 ± 3.42 | -21.21 ± 7.02 | 83 | 97.54 ± 2.88 | -22.36 ± 3.99 | 0.84 |
| PEEP Set (cmH2O) | 20 | 6.95 ± 3.07 | -4.75 ± 24.73 | 59 | 6.47 ± 2.93 | -12.17 ± 35.1 | 0.84 |
| PT (sec) | 22 | 16.04 ± 3.39 | -7.09 ± 25.77 | 82 | 14.67 ± 2.89 | 7.62 ± 35.02 | 0.44 |
| PTT (sec) | 22 | 39.6 ± 20.72 | -8.59 ± 25.11 | 82 | 34.16 ± 11.39 | 6.6 ± 35.26 | 0.63 |
| Phosphorous (mEq/L) | 21 | 3.42 ± 1.13 | -14.47 ± 9.38 | 81 | 3.37 ± 1.09 | -6.77 ± 21.84 | 0.96 |
| Platelets (K/uL) | 22 | 171.27 ± 109.02 | -15.57 ± 6.31 | 83 | 181.0 ± 88.73 | -5.46 ± 36.5 | 0.91 |
| Potassium (mEq/L) | 22 | 4.0 ± 0.58 | -16.44 ± 6.62 | 83 | 4.02 ± 0.5 | -14.1 ± 7.8 | 0.98 |
| RDW (%) | 22 | 16.43 ± 1.86 | -15.21 ± 6.1 | 81 | 15.23 ± 1.84 | -10.95 ± 8.34 | 0.24 |
| Red Blood Cells (m/uL) | 22 | 3.42 ± 0.58 | -15.21 ± 6.1 | 82 | 3.27 ± 0.49 | -10.81 ± 8.37 | 0.65 |
| Respiratory Rate (BPM) | 22 | 22.41 ± 5.28 | -19.85 ± 7.61 | 83 | 20.7 ± 5.63 | -22.4 ± 3.8 | 0.54 |
| Sodium (mEq/L) | 22 | 138.0 ± 4.84 | -17.28 ± 6.79 | 83 | 138.57 ± 4.51 | -13.67 ± 7.85 | 0.87 |
| Temperature C (°C) | 22 | 37.09 ± 0.73 | -19.72 ± 7.01 | 83 | 37.35 ± 0.84 | -20.91 ± 4.76 | 0.51 |
| Total Bilirubin (mg/dL) | 15 | 3.87 ± 4.85 | 29.37 ± 58.6 | 61 | 1.23 ± 1.61 | 30.73 ± 54.06 | 0.37 |
| No. of Previous Cultures (N) | 22 | 1.05 ± 1.56 |  | 83 | 0.35 ± 1.19 |  | 0.39 |
| No. of Previous Resistant Cultures (N) | 22 | 1.36 ± 2.65 |  | 83 | 0.55 ± 3.31 |  | 0.61 |
| White Blood Cells (K/uL) | 22 | 13.46 ± 8.0 | -13.21 ± 10.1 | 83 | 12.83 ± 5.93 | -6.73 ± 21.11 | 0.92 |
| Fraction of fever measurements out of all temperature measurements (%) | 22 | 24 ± 21 |  | 83 | 34 ± 28 |  | 0.39 |
| pCO2 (mmHg) | 20 | 37.15 ± 6.13 | -14.73 ± 16.24 | 75 | 40.83 ± 8.16 | -1.93 ± 40.67 | 0.29 |
| pH (U) (pH) | 20 | 5.68 ± 0.78 | 26.79 ± 69.36 | 72 | 5.68 ± 0.75 | 18.52 ± 40.37 | 0.99 |
| pO2 (mmHg) | 21 | 108.86 ± 43.99 | -14.69 ± 15.81 | 75 | 109.6 ± 35.38 | -0.64 ± 41.49 | 0.98 |

**Table 2.**
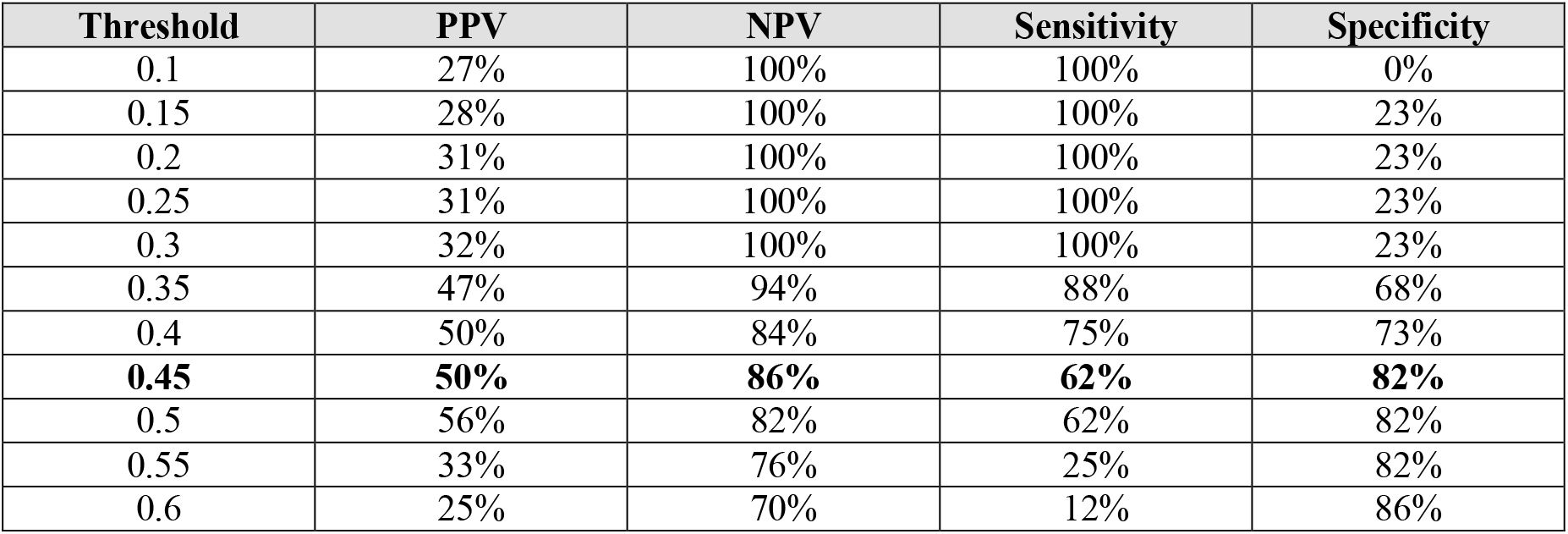
Performance values of the Random Forest DataEnsemble model on the validation set for different risk score thresholds. PPV – Positive Predictive Value, NPV – Negative Predictive Value. Bold – the values for the selected threshold.

**Figure 1.**
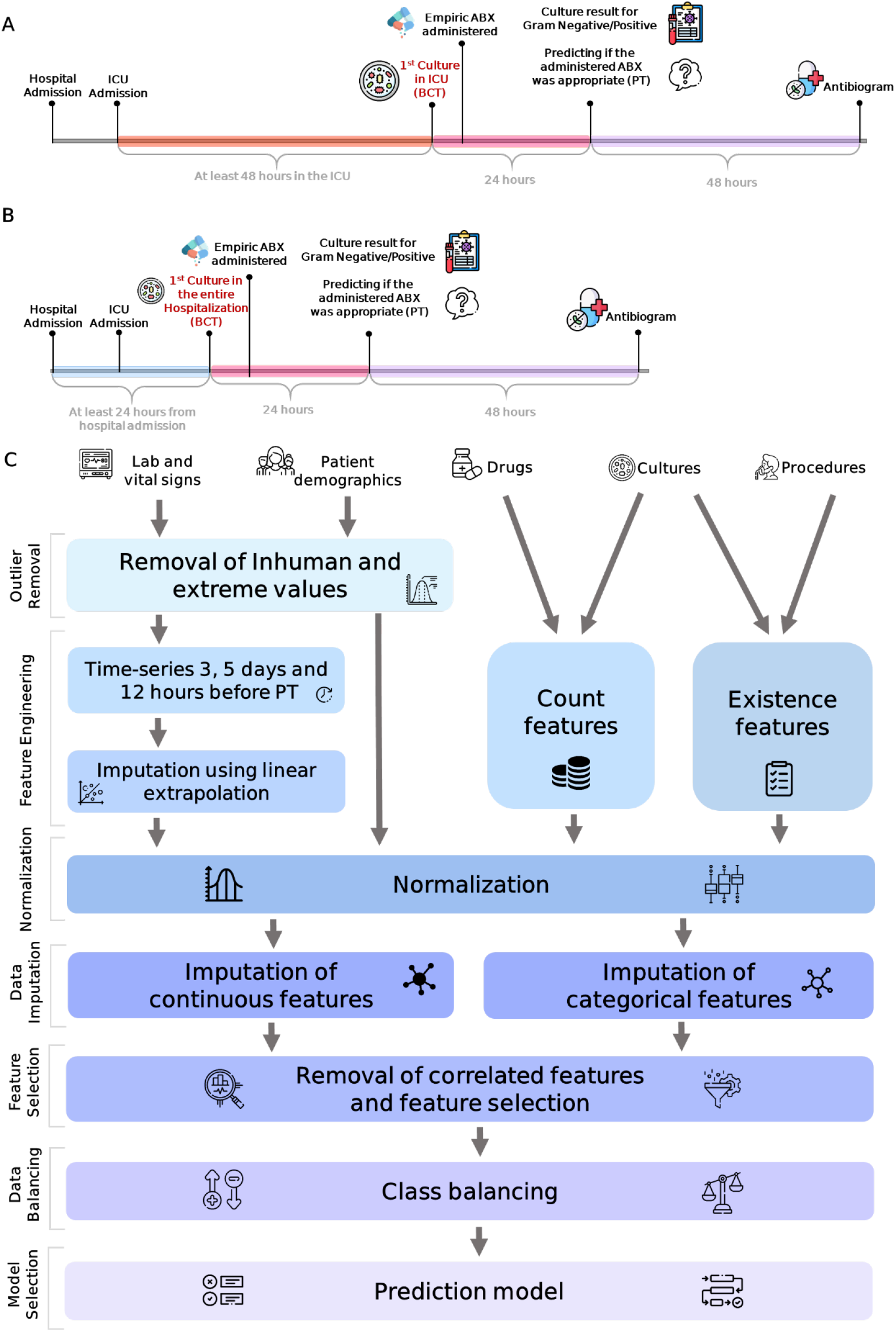
Prediction timeline and the model pipeline. Patients were included in two scenarios: **(A)** At least 48 hours passed from ICU admission until the time the first blood culture taken (BCT) (orange interval). Our model uses also data collected 24 hours after BCT (pink interval) and then returns the prediction whether the antibiotic (ABX) administered was appropriate or not. It takes an additional 48 hours for the antibiogram culture results to return from the lab (purple interval). **(B)** At least 24 hours passed from hospital admission to BCT (light blue interval). Pink and purple intervals are the same as in panel A. **(C) Model pipeline**. Existence features are binary (e.g., existence of a culture resistant to penicillin); Count features are categorical (e.g., count of antibiotic drugs administered to the patient); PT – prediction time.

### Administered Antibiotics Analysis

Analysis of blood culture results and the drugs administered to the patients in our cohort revealed that Coagulase-positive *Staphylococcus aureus* was the most common pathogen, detected in 51% of the patients (69/135). The most common antibiotics administered to patients with that organism were *vancomycin (*50.7%, 35/69) and *levofloxacin* (23%, 16/69). Overall, the most common antibiotic administered to patients was *vancomycin* (57%, 77/135, **Supplementary Figure 1**).

Furthermore, the AST results of those blood cultures revealed the most common pairing of an organism and the antibiotic tested on it. Of the antibiotics tested on Coagulase-positive *Staphylococcus aureus, Gentamicin* had 43 cultures (1/43 had a resistant outcome), *Oxacillin* had 43 (23/43 resistant), and *Levofloxacin* had 42 (25/42 resistant). The most resistant bacteria was *Enterococcus faecium*, which was resistant for at least one type of antibiotic 73% of the times it was observed (38/52), and the antibiotic that had the highest incidence of resistance was *erythromycin*, which experienced resistance 70.8% of the times (34/48) (**Supplementary Figure 2**).

### Model’s Pipeline

In order to develop a robust model that will address the characteristics of our prediction objective, we constructed an extensive pipeline comprised of several steps (**Figure 1C**), and in each step we evaluated a few alternative techniques. We tested each combination of techniques using five iterations of stratified 5-fold cross-validation over the training set and chose the combination that yielded the highest mean AUPR. Below we describe each step briefly. Full details are provided in the Methods section.

The first step in the pipeline is the removal of values that were deemed outliers. We first excluded values that were not in the human range and then removed values based on two different metrics. Afterwards, we filtered out features with missing rate ≥ 30% and removed features with variance ≤ 0.005.

In the next step, we created time-series features utilizing all data points available before PT. We calculated these features using two sets of timeframes, *d* and *d* + 2 days before PT. Missing values in each timeframe were imputed using a linear regression model that was fitted per subject using all the feature values recorded within a larger time-frame, see Data Imputation in Methods. For these features, we evaluated different thresholds for the minimum number of values that are required for the fitting of a linear regression model (*n*) and we evaluated several timeframes (*d* = 2 *and* 3).

The next step was the normalization of the features. We evaluated two approaches: Min-Max scaling and standardization. Then we added a second imputation step to handle missing values that could not be imputed by the linear regression. We used K-Nearest Neighbors (*KNN*) algorithm^19^ with *k* = 5 and tested several distance measures such as Sklearn’s distance method (an Euclidean distance that accounts for missing coordinates), and two new distance measures.

As many of the features were highly correlated, particularly after the addition of the time-series features, we applied two steps of detecting and filtering correlated features. First, we kept only a small number of features derived from the same raw measurement by selecting those with the most significant p-value according to a t-test between the two classes. We tried several numbers of features. Following this step, we filtered highly correlated features based on hierarchical clustering.

After the removal of correlated features, we still had a high-dimensional feature space. Hence, we examined several feature selection methods: (a) Recursive Feature Elimination ^20^, (b) Taking the features with an importance score higher than the model’s mean feature score (e.g., in the logistic regression model, taking the mean beta coefficient), (c) Taking the *K* features with the highest mutual information score^21^ and (d) Taking the *K* features with highest SHAP values^22^. We also tested combinations of these four methods and several possible values of *K*.

Additionally, since our data was imbalanced (roughly 3/4 appropriate and 1/4 inappropriate) we tested the following approaches for oversampling: *ADASYN*^23^, *SMOTENC*^24^, and *BorderlineSMOTE*^25^ with different balancing ratios, and also developed a novel ensemble method for data balancing which we named ‘DataEnsemble’.

The last step was the prediction model selection. Here we evaluated eight different machine learning models: Random Forest^26^, AdaBoost^27^, Logistic Regression^28^, SVM^29^, SGDclassifier^21^, LightGBM^30^, Sklearn’s Gradient Boosting Classifier^21,31^ and Xgboost^32^.

Every combination of techniques applied in each step above was tested in iterated cross validation. The final prediction model chosen was Random Forest DataEnsemble. See Methods for the combination and the parameter values chosen.

### Appropriate Antibiotic Treatment Model

Our model aimed to predict the risk of administering an inappropriate antibiotic treatment to an ICU inpatient. In order to select the optimal model, we used five iterations of stratified 5-fold cross-validation over the training set. In each iteration, we evaluated the model using the mean area under the receiver-operator characteristics curve (AUROC) and area under the precision-recall curve (AUPR) over all five folds. We then averaged these metrics over the five iterations. Each model was evaluated with and without a novel training approach using balanced cohort (‘DataEnsemble’, see Methods). The Random Forest DataEnsemble model had the best performance (**Figure 2 Error! Reference source not found.**) with an AUROC of 82.76±1.46% and an AUPR of 60.61±3.76% on the training set (**Figure 3**). Notably, for seven out of the eight models the DataEnsemble received better median AUPR scores compared to the original model. Thus, Random Forest DataEnsemble was chosen as the final prediction model.

**Figure 2.**
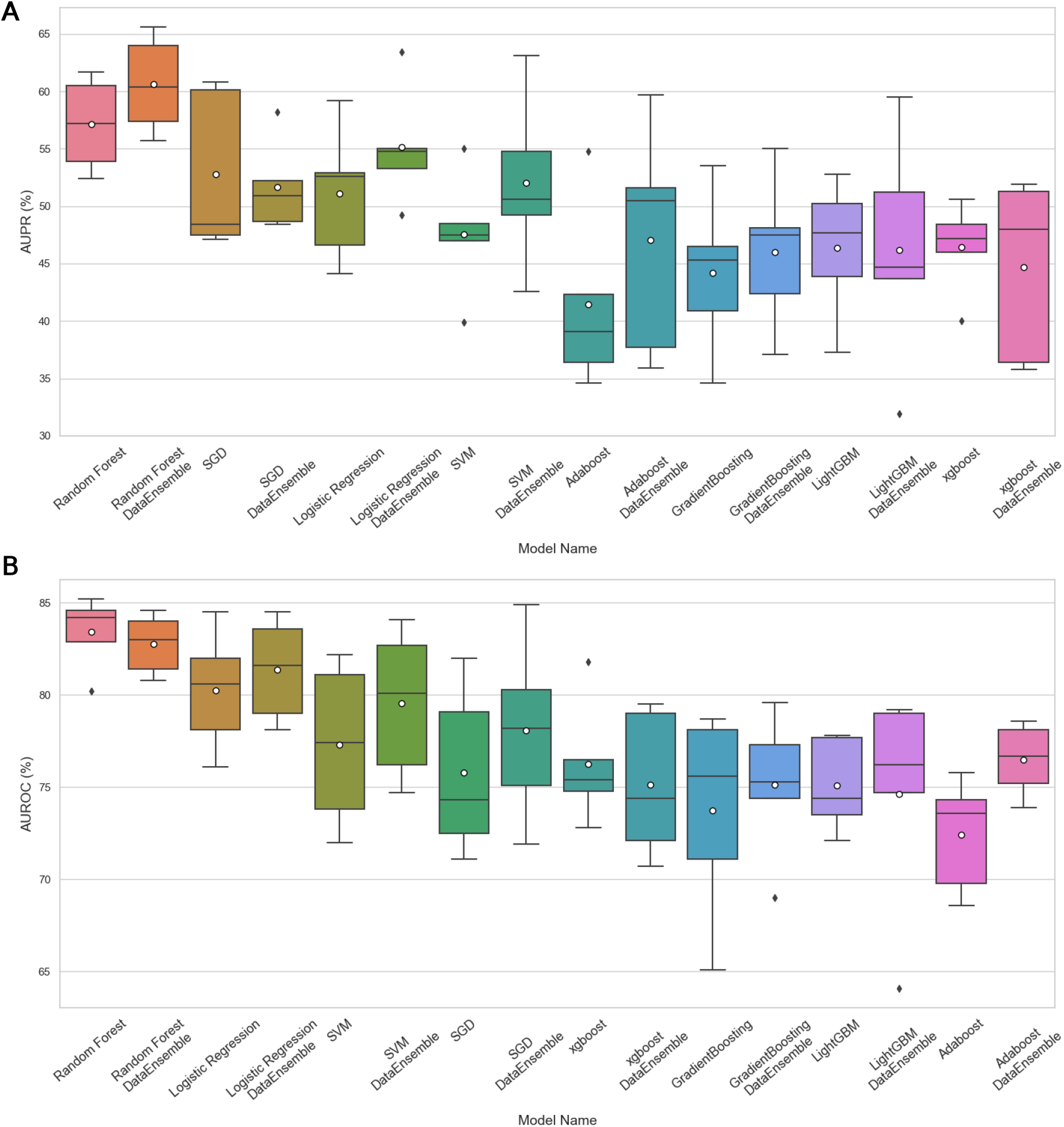
Performance of eight prediction models on the training set. Performance of eight machine learning models with and without the ‘DataEnsemble’ balancing approach for predicting antibiotic appropriateness. Model performance was evaluated using five iterations of 5-fold cross-validation over the training set. The horizontal line indicates the median, the white circle indicates the mean, the box indicates the IQR, the boundaries of the whiskers are the minimum and maximum values, and the black points indicate outliers. **A**. AUPR. **B**. AUROC. The models are sorted by the mean AUPR and AUROC.

**Figure 3.**
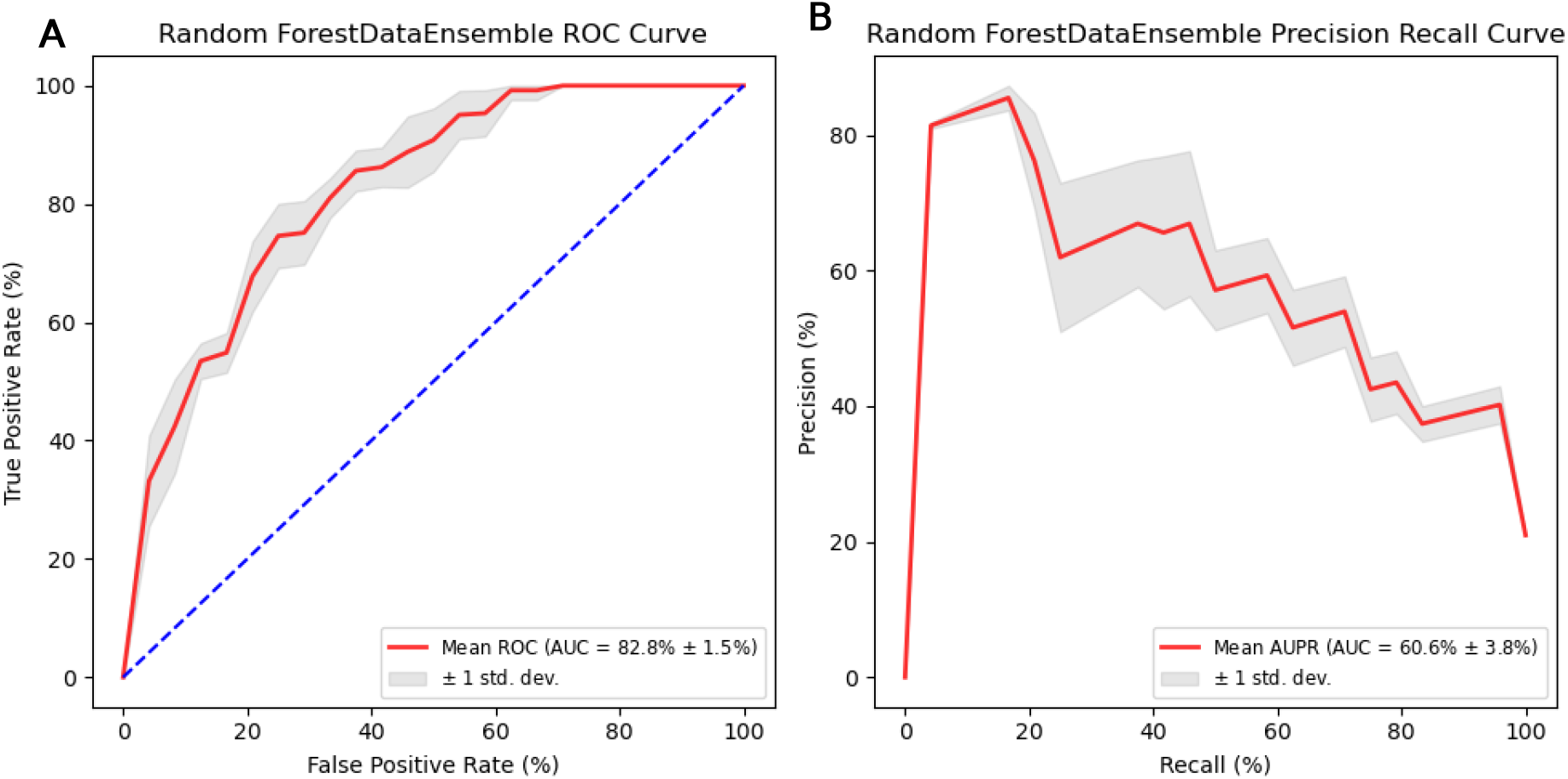
Mean performance of the Random Forest DataEnsemble model on five iterations of 5-fold cross-validation. **A**. AUROC. **B**. AUPR. The red line is the mean, the grey area is ± one standard deviation from the mean.

### Validation

We retrained the Random-Forest model with the selected parameters on the entire training set and applied it on the validation set (**Table 2**, **Figure 4A-B**). A good balance was achieved when using a classification threshold of 0.45 (i.e., classifying all samples with risk score ≥ 0.45 as positive). For that threshold the model achieved a positive predictive value (PPV) of 50%, negative predictive value (NPV) of 86%, sensitivity of 62% and specificity of 82%. In addition, the model achieved an AUROC score of 77.3% and an AUPR score of 40.4%. Those values were lower than those obtained on the training set, however, it is to be expected that a predictor’s performance will be reduced when validated against new samples.

**Figure 4.**
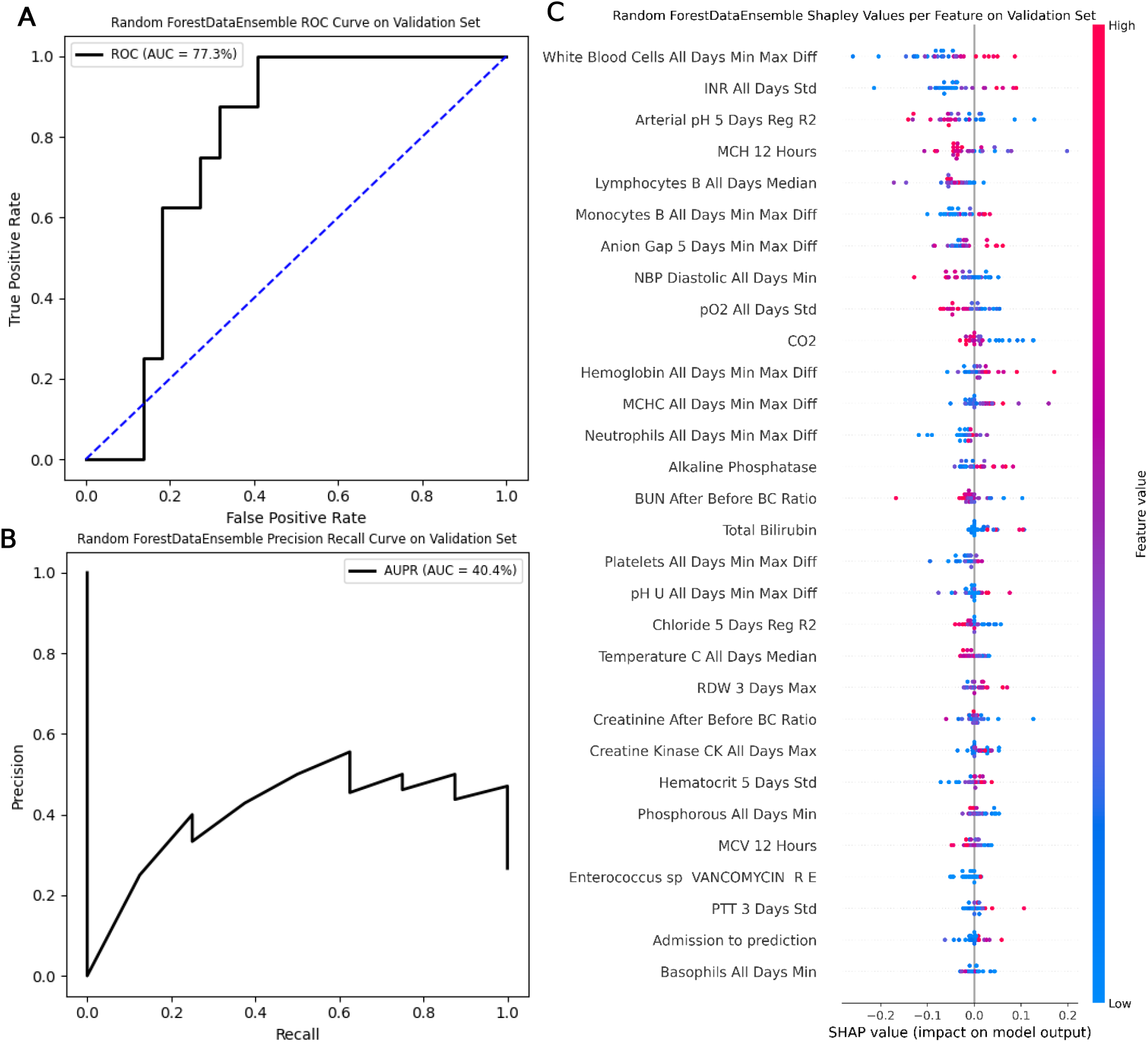
Performance of the Random Forest DataEnsemble model on the validation set. **A**. AUROC, **B**. AUPR. **C**. The thirty features with the highest absolute SHAP values. For each feature the X-axis is the SHAP value, representing the contribution of that value to the model’s decision. The features are ordered in descending mean absolute SHAP values. Each point corresponds to an observation where the color represents the feature value from blue (low value) to red (high value). The sign of the SHAP value indicates whether the feature observation contributes to positive or negative classification. All Days – time-frame of the entire hospitalization up to the prediction time (PT), 3/5 Days – time-frame of 3 or 5 days before PT, 12 Hours – measurement recorded 12 hours prior to PT, Min – minimal value, Max – maximal value, Min Max Diff – difference between the maximal and minimal values measured, Std – standard deviation, Reg R2 - *R*^2^ coefficient of a linear regression model fitted on values in the time-frame, After Before BC Ratio – ratio between the first value recorded after the blood culture was taken and the last value recorded before it, R – resistant culture, E – existence.

Next, we assessed the significance of utilizing data obtained after administration of antibiotics to the patient (i.e, the time between BCT to PT). To accomplish this, the same Random Forest model DataEnsemble was applied using only data obtained prior to BCT. The resulting model exhibited poor performance, achieving AUROC 58% and AUPR 29.8%, which mirrors the proportion of positive samples in the validation set at 26.67%. Subsequently, the pipeline parameters were optimized for the best mean AUPR on the training set and the model was evaluated on the validation set. The results were similar, yielding AUROC 55.1% and AUPR 27.7%. These findings show the importance of using data from the period following the drug administration to the patient. It is evident that training the model solely on pre-culture data without also using the data after the drug intervention results in near-random predictions, as the model lacks sufficient informative values.

We also wished to assess the contribution of data obtained before antibiotic administration to the prediction. For this goal, we applied the same Random Forest DataEnsemble model using only data obtained after the blood culture was taken (i.e., from BCT to PT). Again, the resulting model exhibited poor performance, achieving AUROC 61.4% and AUPR 31%. Optimizing the pipeline parameters for the best mean AUPR on the training set and evaluating the model on the validation set resulted in similar performance, yielding AUROC 60.2% and AUPR 30.1%. Hence, relying solely on post-culture or pre-culture data for training the model leads to poor predictions, and incorporating information both from before and after drug administration greatly improves prediction quality.

### Feature Importance

Analysis of the features created for our model showed that none of the raw lab measurements and vital signs measurement were significant discriminators. However, previous cultures and especially resistant cultures were significantly associated with the inappropriate class (**Supplementary Table 3**). Moreover, the existence of any Ascites lab test is also associated with the inappropriate classification.

The contribution of each feature to the model’s risk score is estimated using SHAP values^22^ (**Figure 4C**). Most of the features that had a substantial impact on the model were time-series features of vital signs and lab measurements. The most important features of the model were the difference between maximum and minimum white blood cell count (WBC) measured during the hospitalization, the standard deviation of INR values measured during the entire hospitalization, *R*^2^ of a regression model of arterial pH values in the 5-day timeframe before PT, and the mean corpuscular hemoglobin (MCH) measured 12 hours before PT. Although total WBC count is a common laboratory marker for identifying patients with high risk for bacterial infection (BI), studies have shown that WBC count had only minor discriminatory power in identifying patients with BI^33–35^.

## Discussion

Approximately 70% of patients admitted to the ICU receive antibiotic treatment^36^. However, the percentage of patients who do not receive adequate therapy within the first 24 hours of a bloodstream infection (BSI) is alarmingly high, reaching 47%^37^. On the other hand, ill-advised and excessive antibiotic use can contribute to the global antibiotic resistance problem^4^. In this study, we propose a machine learning algorithm to predict inappropriate empiric antibiotic treatment in patients with ICU-acquired bacteremia. Previous research has focused on utilization of machine learning models for early prediction of ICU-acquired BSI^38^, outcomes of BSI^39^, and antibiotic resistance in BSI^40,41^ and urinary tract infections^42,43^. Studies^39,40^ predicted antibiotic susceptibility by creating a specific model for each antibiotic type. In contrast, the problem of antibiotic treatment appropriateness is not concerned with the resistance to each type of antibiotic, but evaluates whether the treatment administered was effective by assessing the patient’s response to it. Due to the limited size of the available cohort, the model described in this study was not specifically trained for individual antibiotic types. Consequently, we developed one general model for predicting the appropriateness of the antibiotic treatment, without the need to specify which antibiotic was administered to the patient. The purpose of this model is to discern the physiological response to an appropriate antibiotic treatment from that of an inappropriate treatment. To the best of our knowledge, no prior studies have addressed the problem of determining the appropriateness of antibiotic treatment.

Our algorithm demonstrated promising performance both in cross-validation and in validation of an independent sample of patients (with AUROC scores of 82.76% and 77.27%, and AUPR scores of 60.61%, and 40.44%, respectively). These results suggest that with the use of readily accessible EMR data, it is possible to predict the appropriateness of an antibiotic treatment 48 hours before the full antibiogram results are available and assist in the clinical assessment of the patient. The substantial reduction in mismatched treatment facilitated by machine learning-based recommendations that take into account the patient’s medical history and records can pave the way for a future framework in which clinicians will routinely consult such algorithms and adjust the antibiotic treatment of patients accordingly. Adoption of the model in clinical practice could lead to a machine learning-guided personalized antibiotic prescription and help reduce treatment failure and overall use of antibiotics, contributing to the global effort to combat antibiotic resistance.

The models’ prediction was primarily driven by the patterns in the time-series features, such as the difference in the median measurement of WBC collected in the 5-day and 3-day time-frames prior to PT (**Figure 4C**). Some of these features were previously studied in relation to BI and were recognized as significantly associated with it. However, the temporal behavior of most of these features was not checked in relation to BI. Moreover, to the best of our knowledge, no study identified clinical measurements that are most relevant to predicting antibiotic treatment appropriateness. Notably, no raw measurement by itself was statistically significant for discriminating between appropriate and inappropriate treatments. Therefore, examining the features selected by our machine learning model can provide valuable insight into such discrimination. By identifying the predictors with the highest impact on the model’s outcome, doctors can focus on those lab measurements and vital signs.

In addition to time series features, our model utilized known risk factors for antibiotic resistant infections as features, such as previous antibiotic resistant infections, antibiotic that were previously administered, invasive procedures and culture sample sites^44–46^. Many of these features were also shown as predictive for antibiotic resistance in machine learning models that used EMR data^42,47,48^.

In this study, we chose to set PT to 24 hours after the blood culture was taken, as the results of the gram-staining are typically retrieved at this time^9^, and thus at that time clinicians could make adjustments to the patient’s antibiotic treatment. Providing additional information at this time can improve decision-making by the doctors, potentially affecting the patient’s outcome. Moreover, our cohort included only ICU inpatients with microbiological confirmation of a bacterial infection. However, studies have shown that only about 19.5% of inpatients with bacteremia have a positive blood culture^49^. Therefore, it is plausible that many of the patients with bacteremia will potentially not be considered for our model. Developing a model where PT is instead set to when gram stain results are already available, and the microbiological confirmation of the bacteria exists, can increase the percentage of relevant inpatients considered by the model. Furthermore, our findings demonstrate that data collected during the additional 24 hours lead to a significantly better prediction in comparison to a model trained solely on data obtained prior to the blood culture.

Our study has several limitations. First, in our study we filtered out contaminants, while they might be considered eligible cultures for our prediction since no information is provided to classify them as contaminants at the time of gram stain. Additionally, our model was trained and validated on a relatively small dataset from one medical center, and should be tested on data from other medical centers. Finally, conducting a prospective evaluation is necessary to assess the model performance in practical scenarios.

It is also important to note that the data collected per patient only pertained to the hospitalization during which the blood culture was taken. Future studies could benefit from incorporating the complete medical history and previous hospitalization records of a patient^42^. In particular, the use of data on previous cultures can enhance the model’s predictive ability, as previous instances of recurrent infections are associated with a higher risk of resistant infection in subsequent hospitalizations^50^.

In addition to the contribution to predicting antibiotic resistance, this study also proposes a new pipeline for medical decision support. It outlines techniques to address challenges commonly encountered in EMRs, such as limited and imbalanced datasets and high rates of missing values. The key methods described here can serve as starting points for such an approach, but the specific model, parameters, and feature extraction process should be tailored to the medical question and the data.

## Methods

### Inclusion and Exclusion Criteria

All patients admitted directly to the emergency department or ICU who had blood cultures that were not contaminated (i.e., blood culture results of Coagulase-negative *Staphylococcus, Diphtheroids, Bacillus, Aerococcus viridans, Aerococcus, Propionibacterium, Viridans streptococci, Lactobacillus*, and *Staphylococcus epidermidis*) or were not canceled during their hospitalizations were considered for this study’s cohort. Out of those, to identify patients with hospital acquired infection we included only patients who satisfied at least one of the following conditions: (a) they were hospitalized for at least 48 hours in the ICU and had their first blood culture in the ICU collected there after that time. Only the first culture collected in the ICU was used for labeling. (b) their first culture in the entire hospitalization was collected in the ICU and at least 24 hours after hospital admission (Error! Reference source not found.).

### The Cohort

We used the MIMIC-III database, containing data of 38,597 distinct adult patients^18^. Our exclusion criteria resulted in a total of 135 patients, who were split into training and validation sets. Our training set included EMRs of 105 inpatients, of whom 83 received appropriate antibiotic treatment and 22 received inappropriate antibiotic treatment. The validation set included 30 inpatients of whom 22 received appropriate treatment and 8 received inappropriate treatment.

### Outcome Definition

Microbiological cultures are routinely drawn in ICU. We defined the blood culture time (BCT) as the time of the culture sampling, and PT as 24 hours after culture time. Only records charted before PT were used by the model.

Antibiogram results are usually available within 72 hours of culture sampling^9^, so prediction after 24 hours may allow the physician to reconsider the antibiotic empirical treatment 48 hours before the antibiogram results. The 24-hour window enables one to obtain features that help assess the response of different clinical measures to the empiric antibiotic treatment (for example, the ratio between white blood cells levels before and after the empiric antibiotic treatment).

Patient treatments were designated as appropriate (negative class) or inappropriate (positive class) treatment based on the results of the culture, AST and the empirical antibiotic that was administered. The inappropriate class was defined as an antibiotic treatment where the pathogen was either not affected by the antibiotic or resistant to it. Appropriateness was decided by an internal medicine specialist and an infectious disease specialist who reviewed together the antibiotics administered and the antibiogram results for each patient.

### Outlier Removal

#### Inhuman Values

To eliminate measurements that were grossly incorrect due to manual typos or technical errors, we manually defined with clinicians a range of possible values per each feature (including pathological values), and excluded values outside this range. A total of 711 values (0.4% of the values of all features) were excluded in this step, see **Supplementary Table 4**.

#### Extreme Values

For the remaining values, we checked two approaches to removing extreme measurements. Both of these methods were calculated on the training set, and were later applied on the validation set. The first method used the IQR. Denote by *q*_0.75_ (*q*_0.25_) the value at the 75^th^ (25^th^) percentile and set *q*_*diff*_ = 1.5 × (*q*_0.75_ − *q*_0.25_). Then only values in the range *q*_0.25_ − *q*_*diff*_ < *x* < *q*_0.75_ + *q*_*diff*_ were kept.

The second approach used Z-scores, filtering out values that are more than two standard deviations from the mean of the feature.

We analyzed the percentage of values that were removed after applying both methods. Z-score discarded a mean of 4.12% of the feature values and a median of 3.69%, while IQR removed a mean of 5.44% and a median of 3.67% (**Supplementary Table 4**). Following these results, we used the Z-score approach.

### Normalization

We evaluated two approaches for feature normalization. The first is the normalization of all features to values between 0 and 1 according to the maximum (*X*_*max*_) and minimum (*X*_*min*_) values of each feature in the training data set.

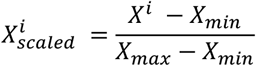

The second was standardization to a normal distribution with a mean of zero and a standard deviation equal to one.

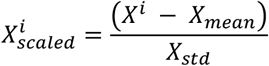

Both normalizations were fitted on the training set data, and later applied on the validation set as well. The normalization method chosen was standardization as it yielded better results.

### Feature Engineering

The features created for the model are composed of six main categories: (1) patient demographics, (2) lab measurements, (3) vital signs, (4) drug administration, (5) previous lab cultures, (6) medical procedures. We tested removal of features with high missing rates, for rates 20%, 30%, 40%, and 50%, and chose to exclude features with missing rate > 30%. Moreover, after the feature engineering process (see below), features with variance < 0.005 were excluded as well.

#### Demographics

The demographic features included, among others, age, gender, and ethnicity, as well as time since admission to the hospital and to the ICU, and measurements such as weight and BMI.

#### Lab measurements and vital signs

We used as features the median, standard deviation, minimal value (min), maximal value (max), and their difference (min-max diff) per each time-frame described above. See **Supplementary text** for more details.

#### Drugs

We mapped all the drugs into 11 clinically relevant groups (**Supplementary Table 2**) with the help of a general physician. For each drug group, and each of the time-frames described above, we collected the total number of drugs from the group that the patient received.

#### Cultures

We extracted binary features indicating the properties of previous culture taken from the patient, when available. See **Supplementary text** for more details.

#### Medical procedures

Finally, we added binary features for four categories of invasive procedures that frequently cause infection: Arterial Line, Catheter, Ventilation, and Tubes (**Supplementary Table 5**), and indicated if the patient has undergone a procedure from each category.

The mean time from the first lab or vital sign measurement to PT was 7.15±4.65 days (**Supplementary Figure 3**). Therefore, we generated time-series features for lab measurements, vital signs, and drugs for two time-frames: *d* and *d* + 2 days before PT. We tested *d* = 3 and 4, and 3 yielded better results. Additionally, for lab measurements and vital signs, we also used a time-frame of the entire hospitalization period up to PT. See **Supplementary text** for more details.

### Data Imputation

Missing values were observed mainly in lab measurements and vital signs. For repeatedly measured values, a linear regression model was fitted (see ‘Feature engineering’). We imputed the missing values of features in a certain timeframe based on those linear regression models. This strategy assumed that missing values are more accurately imputed using patient-specific measurements rather than values of all patients. Regression was performed per 3 or 5-day time-frame. If a patient was missing max, min, median, or min-max-diff time features in a certain time-frame, we extended the time-frame used to impute these values to 5 and 10 days, respectively. Moreover, the feature value 12 hours before PT was imputed using the 3-day linear regression, and if a regression model was not available for this time-frame, 5-day linear regression was used. Since large regression coefficients can lead to extreme imputed values, all the values produced by this extrapolation method underwent extreme and non-human values removal (**Figure 5**).

**Figure 5.**
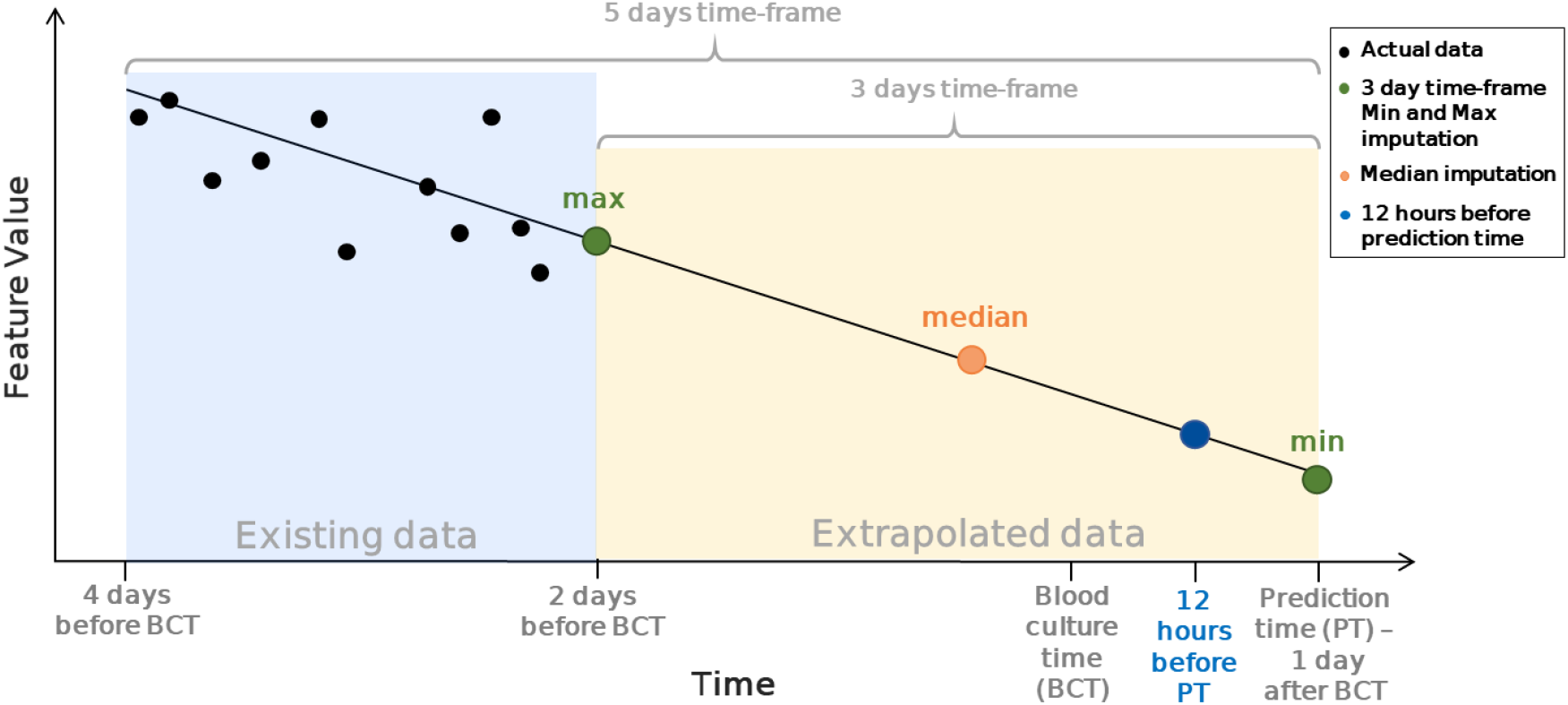
Illustration of the imputation scheme for time-series features. For a patient with missing data in the 3-day time-frame (yellow), the values of max, median, min, and at 12 hours before prediction time are imputed using linear regression calculated based on existing values (black dots) in the 2 days before the beginning of the time-frame (light blue).

The rest of the time-series features, other continuous features (e.g. last lab measurement recorded), and instances where there were not enough values for the fitting of a linear regression model (see ‘Feature Engineering‘), were imputed based on the *KNN* algorithm^19^ with *k* = 5. In order to prevent vectors with high missing rate from being considered “closer” to all the other vectors, we developed two distance methods in addition to Sklearn’s weighted distance metric and evaluated them to choose the best one (see Supplementary Information).

### Removal of Correlated Features

The creation of multiple time-series features in different time frames, as well as the collection of a variety of lab measurements and vital signs that reflect the same trends in patients’ medical condition, created feature redundancy. Two different methods were developed to deal with this problem, using clustering.

In the first method we clustered features based on the type of original measurement they were derived from (e.g., all time-series features derived from heart rate measurements) and filtered only *n*_*keep*_ features from each cluster that had the best p-value for association with the target (*n*_*keep*_ ∈ [1,2,3,4,5]).

In the second method we filtered out features with high correlation to other features. A correlation matrix *C* of all the features was created and transformed into a distance matrix *M*_*ij*_ = 1 − *C*_*ij*_. This matrix *M* was then used for hierarchical clustering in which the final clusters were formed such that no two features in the cluster had a cophenetic distance greater than 1 minus a correlation threshold. The correlation thresholds 0.55, 0.6, 0.65, 0.7, 0.75, 0.8 were tested and 0.7 was chosen. Out of each cluster, only the feature with the best p-value for association with the target was kept. After comparing the effect of those parameters on the model’s performance, we kept only one feature per each of the raw features (*n*_*keep*_ = 1).

### Feature Selection

Four methods of feature selection were evaluated. The first method is Recursive Feature Elimination with Cross Validation (RFECV) ^20,21^. The second method utilizes the model’s default feature importance method and selects only features with importance higher than the mean importance of all features. The third method is filtration of *K* features with the best Shap values^22^. The fourth method selects the *K* features with the highest mutual information score with the target. The *K* values 20, 25, 30, 35, 40, 45, 50 were evaluated for those two latter methods, and the best value *K* = 45 was selected.

In order to increase the robustness of feature selection, we summarized the results of the four feature selection methods tested in two ways. First, we checked the union of all the features selected by the methods. Second, we chose only the features that were selected by at least two of the four methods. The union of the features selected by all four methods using yielded the best results.

### Model Development

#### Data Balancing

Three different methods for oversampling were tested. The first two methods, *ADASYN*^23^, and *BorderlineSMOTE*^25^ generate synthetic data mostly based on the “most difficult” samples for learning, and they both assume that all features are continuous. The third method, *SMOTENC*^24^ distinguishes between continuous and categorical features and samples those features accordingly. Moreover, for each method, we tested different balancing ratios for the inappropriate treatment class, which was the smaller class, taking ratios of 0.3, 0.35, 0.4, 0.45 and 0.5.

Moreover, we developed an ensemble model (*DataEnsemble*) that is composed of two instances of the same model trained on all positive samples and a different, disjoint subset of negative samples. Therefore, each model in the ensemble is trained on a proportion of 1:2 for positive compared to negative patients. The risk score of this model is the average score of the two models in the ensemble (**Figure 6 Error! Reference source not found.**). After evaluating all those methods, *BorderlineSMOTE* with a balancing ratio of 0.3 and utilization of DataEnsemble model were chosen for data balancing.

**Figure 6.**
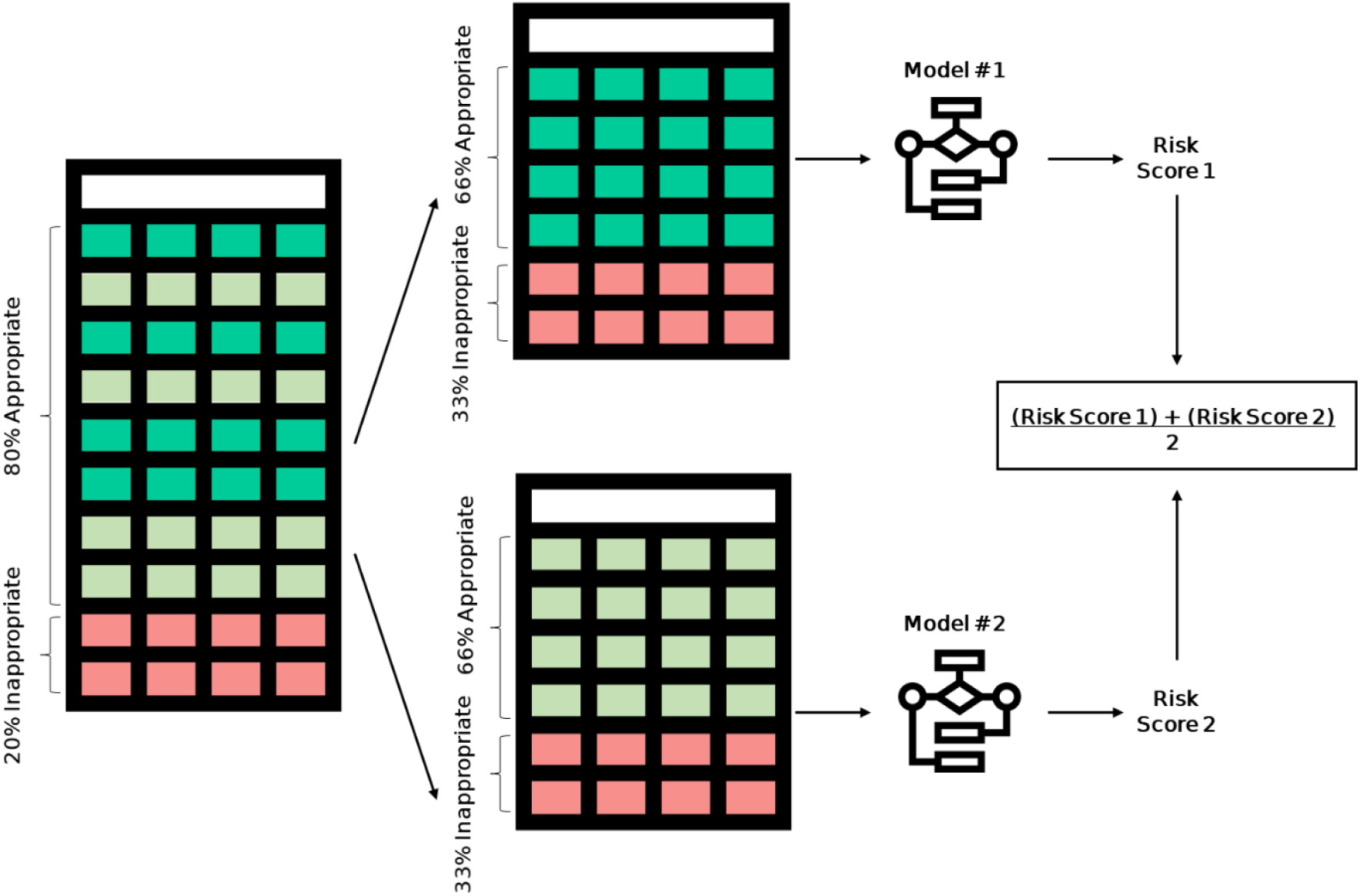
Depiction of the “DataEnsemble”. On the left is the original dataset, rows of inpatients that received an appropriate antibiotic treatment (negatives) are colored in shades of green, and rows of inpatients that had received inappropriate treatment (positives) are colored in red. On the right are two subsets of the data, each containing all the positive patients, and a random, disjoint subset of the negative patients. The “DataEnsemble” is composed of identical models, each trained on a different subset of the data.

Another possible way to handle class imbalance is using class weights. Class weights adjust the loss function of the model to penalize the misclassification of the minority more heavily than those of the majority class, thus improving the model’s learning process on the minority class. We evaluated different forms to allocate a high weight to the positive class (**Supplementary Table 7**), but did not obtain any substantial enhancement in the performance of the model.

#### Model Selection

In order to choose the best model possible for our data, eight different binary classification models were compared - Random Forest^26^, AdaBoost^27^, Logistic Regression^28^, SVM^29^, SGDclassifier^21^, LightGBM^30^, sklearn’s Gradient Boosting Classifier^21,31^ and Xgboost^32^. For each model we created a DataEnsemble model as described above.

#### Hyperparameter Optimization

After choosing the best model and pipeline parameters using an exhaustive search over the parameter combinations (e.g., data normalization method, see Model’s Pipeline), we used grid search to evaluate the effect of different model hyperparameters (e.g., Random Forest’s max depth) on the results of the model trained on each of the five iterations of 5-fold cross validation on the training set. We tried different parameter combinations (**Supplementary Table 6**) and chose the combination that yielded the best mean AUPR results.

## Data Availability

The MIMIC-III database analyzed in this study is available on PhysioNet repository^51^.

## Code Availability

The code used for data processing and model development is available at https://github.com/Shamir-Lab/ABXAppropriatenessML.

## Acknowledgements

We thank Sarah Amar, MD for helpful inputs. Study supported in part by the Israel Science Foundation (grant No. 3165/19, within the Israel Precision Medicine Partnership program, and grant No. 2206/22) and by the Tel Aviv University Center for AI and Data Science (TAD). EG, ER and DC are supported in part by fellowships from the Edmond J. Safra Center for Bioinformatics at Tel-Aviv University. This work was carried out in partial fulfillment of the requirements for the Ph.D. degree of D.C. at the Blavatnik School of Computer Science, Tel Aviv University. All icons used in this paper are designed by Freepik and are available at https://www.flaticon.com/. The funders had no role in study design, data collection and analysis, decision to publish, or preparation of the manuscript.

## Author Information

These authors contributed equally: Ella Goldschmidt and Ella Rannon.

These authors contributed equally: Dan Coster and Ron Shamir.

## Contributions

E.G., E.R., D.C., R.S. conceived and designed the analysis; E.G., E.R. performed the data analysis, model development and model evaluation; E.G., E.R., D.C., R.S. contributed to the study design; A.W., D.B. assisted in the evaluation of the clinical aspects and data interpretation; E.G., E.R., D.C., R.S. wrote the manuscript.

## Supplementary Information

**Supplementary Figure 1.**
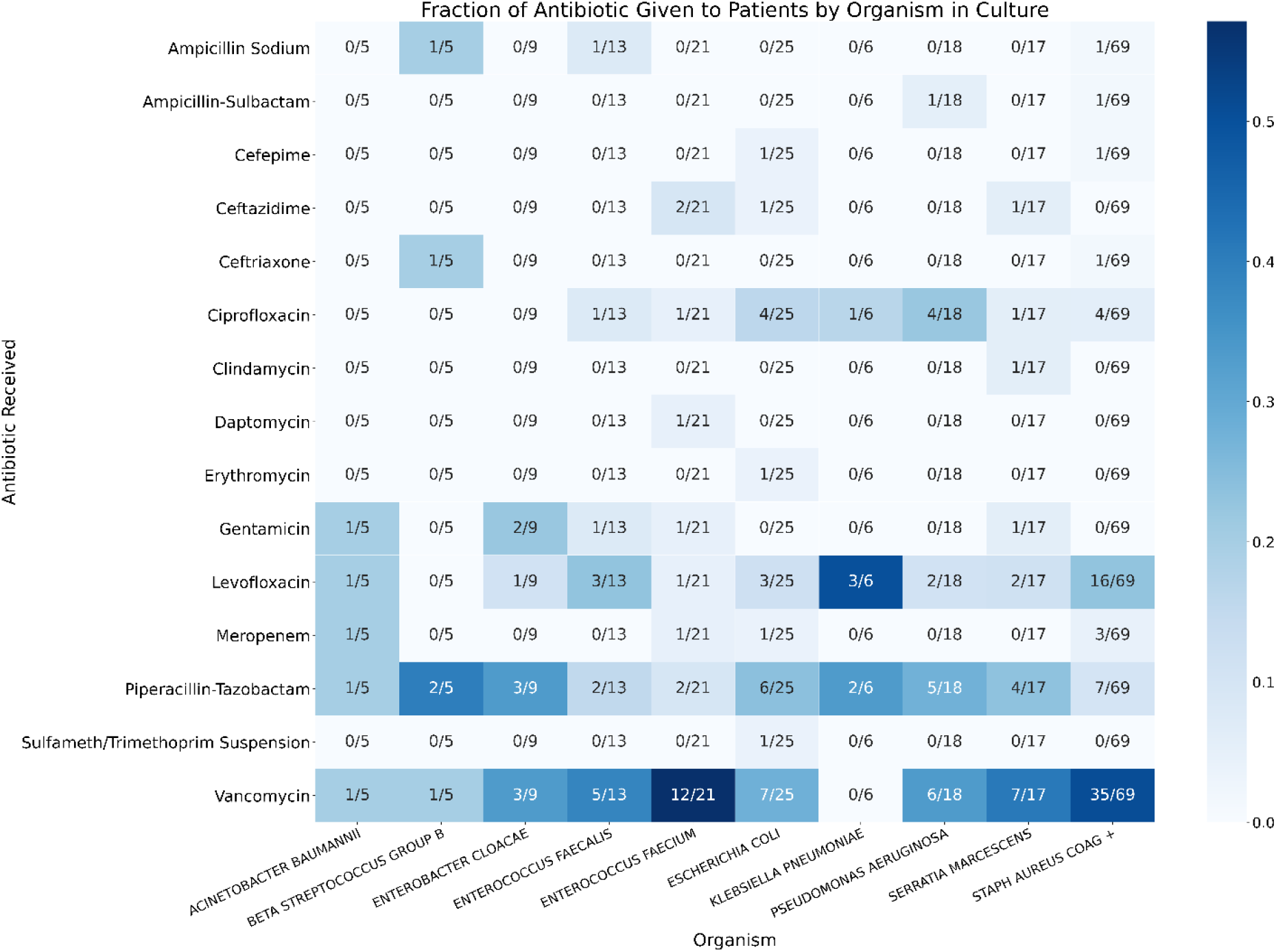
Antibiotics administered to ICU inpatients by the organism detected in their blood culture. The X-axis is the organism; the y-axis is the antibiotic administered to patients. Each cell contains a fraction of patients that received the specific antibiotic out of all patients that had the organism detected in their culture. Color represents the value of the fraction from light blue (low value) to dark blue (high value). Both rows and columns were filtered to present only antibiotics or organisms that appeared in at least four patients. This analysis takes into account all blood cultures taken until the blood culture time used by our model.

**Supplementary Figure 2.**
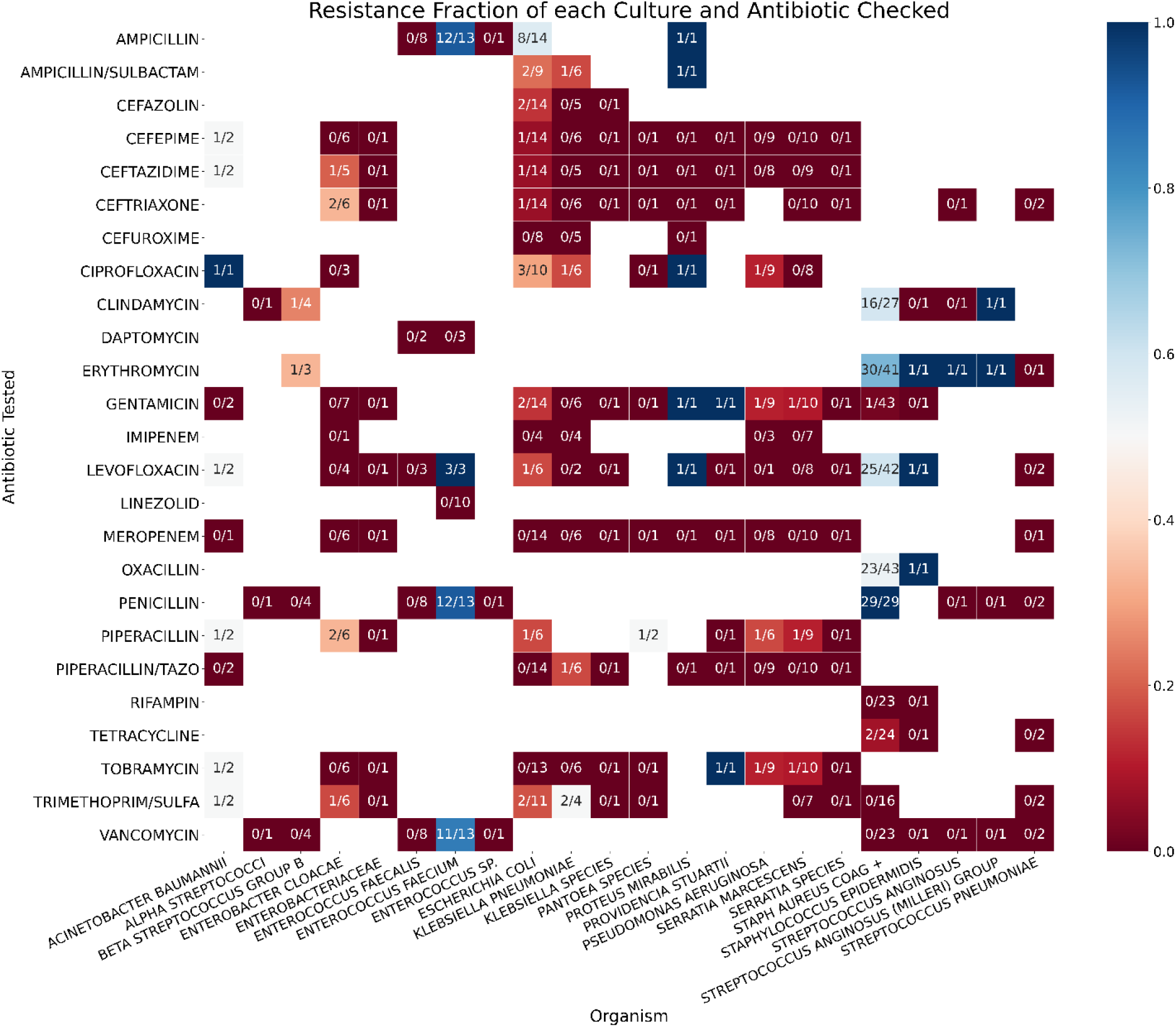
Antibiotic Resistance for each organism and antibiotic checked. The X-axis is the organism; the y-axis is the antibiotic tested. Each cell contains the fraction of patients that had a bacteria that is resistant to that specific antibiotic out of all patients that had that bacteria and had that antibiotic administered to them. Color represents the value of the fraction from dark red (low value) to dark blue (high value). Rows were filtered to contain only antibiotics that were tested on cultures of at least four patients. This analysis takes into account all blood cultures taken until the blood culture time used by our model.

**Supplementary Figure 3.**
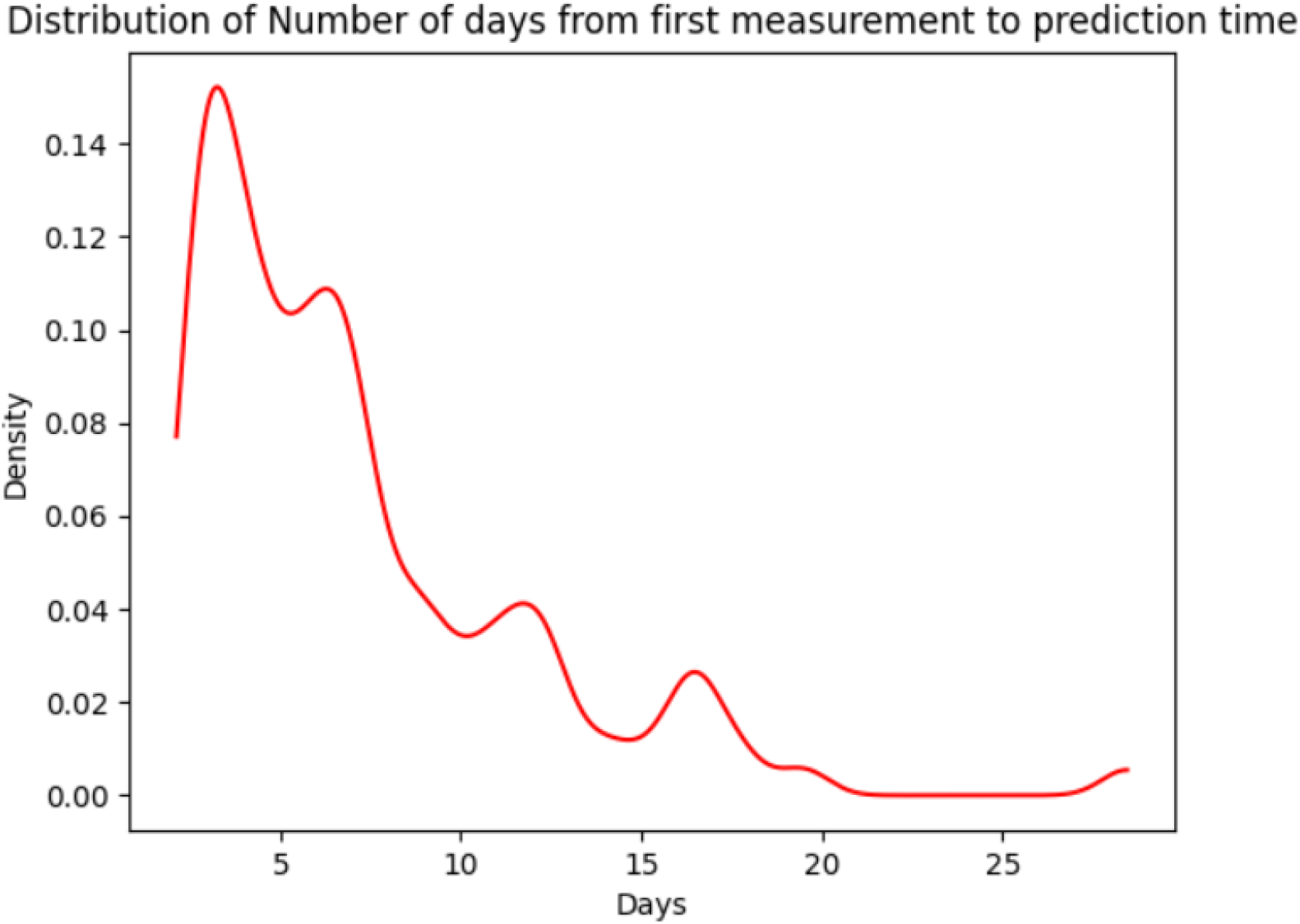
Distribution of number of days from patients’ first measurement to prediction time. The distribution is calculated across all the patients in our data set taking into account all the patients’ vital signs and lab measurements.

**Supplementary Table 1.** Statistics of Drugs administered in the cohort. Each drug feature represents the count of all the drugs of this category that were administered before the prediction time per subject. We used student’s t-test to compare between the groups.

| Feature | Inappropriate |  | Appropriate |  | P-value |
| --- | --- | --- | --- | --- | --- |
| | N | Mean ± SD | N | Mean ± SD | |
| Anesthesia and pain treatment | 22 | 7.68 ± 16.68 | 83 | 8.95 ± 27.82 | 0.93 |
| Antibiotics | 22 | 3.23 ± 8.62 | 83 | 1.4 ± 4.45 | 0.71 |
| Cardiovascular | 22 | 5.91 ± 15.82 | 83 | 11.72 ± 36.18 | 0.64 |
| Insulin | 22 | 5.14 ± 19.27 | 83 | 2.69 ± 10.0 | 0.86 |
| Infusion nutrition | 22 | 30.18 ± 88.85 | 83 | 21.4 ± 56.83 | 0.9 |
| Proton Pump Inhibition (PPI) | 22 | 1.0 ± 2.31 | 83 | 1.07 ± 2.49 | 0.97 |
| Coagulation | 22 | 2.77 ± 7.03 | 83 | 1.72 ± 5.95 | 0.83 |

**Supplementary Table 2.** Mapping of MIMIC-III “itemid”s to the 11 drug categories used in our model.

| Drug category | itemids |
| --- | --- |
| Infusion nutrition | 220862, 220864, 227525, 225823, 225941, 225827, 225825, 225795, 220950, 228140, 228141, 228142, 220949, 220952, 225174, 225167, 227536, 225828, 225159, 225158, 228341, 225161, 225166, 220995, 227533 |
| Coagulation | 221261, 221319, 225147, 225148, 225913, 225171, 221689, 225906, 225151, 225914, 221733, 225908, 220970, 225152, 225975, 221892, 225168, 227532, 225170, 225157, 227535 |
| Swallow nutritional supplement | 225948, 225970, 227977, 227976, 227978, 227979, 225947, 227091, 226875, 225937, 226877, 227698, 227699, 227696, 227695, 225832, 228356, 228359, 226023, 226020, 226021, 226022, 221207, 226027, 226024, 226026, 225928, 228131, 228132, 228133, 228134, 228135, 225834, 225969, 225801, 225995, 225996, 225994, 225830, 225835, 228348, 228351, 227973, 227974, 227975, 226019, 226016, 226017, 227518, 225931, 226882, 226881, 226880, 226031, 226028, 226030, 221036, 225993, 226039, 226036, 226038, 225930, 228383, 227370, 225920, 225929, 225915, 228361, 228363, 226047, 226044, 226045, 226046, 225935, 226051, 226048, 226049, 226050, 225936, 225991, 225833, 225916, 225917, 228364, 228367, 226059, 226058, 225934 |
| Cardiovascular | 221282, 221347, 228339, 221393, 221456, 221653, 221662, 221289, 221429, 221794, 228340, 221828, 227692, 227522, 225153, 222011, 227523, 227524, 227531, 225974, 221986, 222037, 222042, 222056, 222051, 221906, 221749, 222151, 222190, 222315, 222318 |
| Anesthesia and pain treatment | 228315, 221555, 225150, 221623, 221468, 221744, 225942, 221833, 221712, 225945, 221385, 225973, 227520, 221668, 225154, 222021, 225156, 222168, 222062 |
| Insulin | 223257, 223260, 223262, 223261, 223259, 223258, 225155 |
| Antibiotic | 225840, 225842, 225843, 225845, 225847, 225899, 225850, 225851, 225853, 225855, 225859, 225860, 225862, 227534, 225863, 225865, 225866, 225868, 225875, 225876, 225877, 227691, 225879, 225881, 225883, 225884, 225886, 225888, 225889, 225890, 225892, 225893, 225895, 225898, 225902, 225798 |
| Antifungal | 225838, 225848, 225869, 225885, 225905 |
| PPI (proton pump inhibition) | 225912, 225907, 225909, 227694, 225910, 225911 |
| Asthma | 221342 |
| Epilepsy | 227689, 227690, 228316 |

**Supplementary Table 3.**
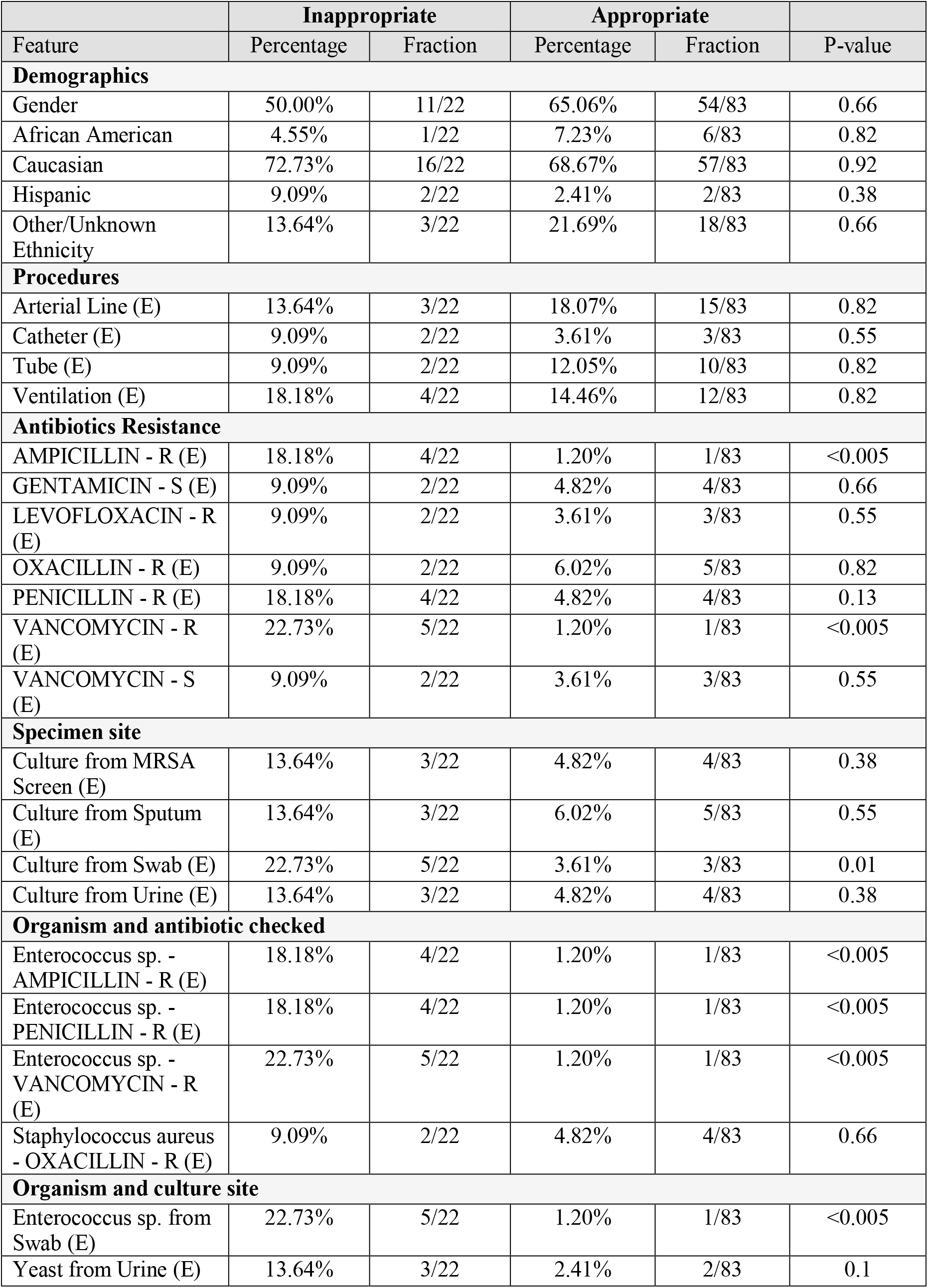
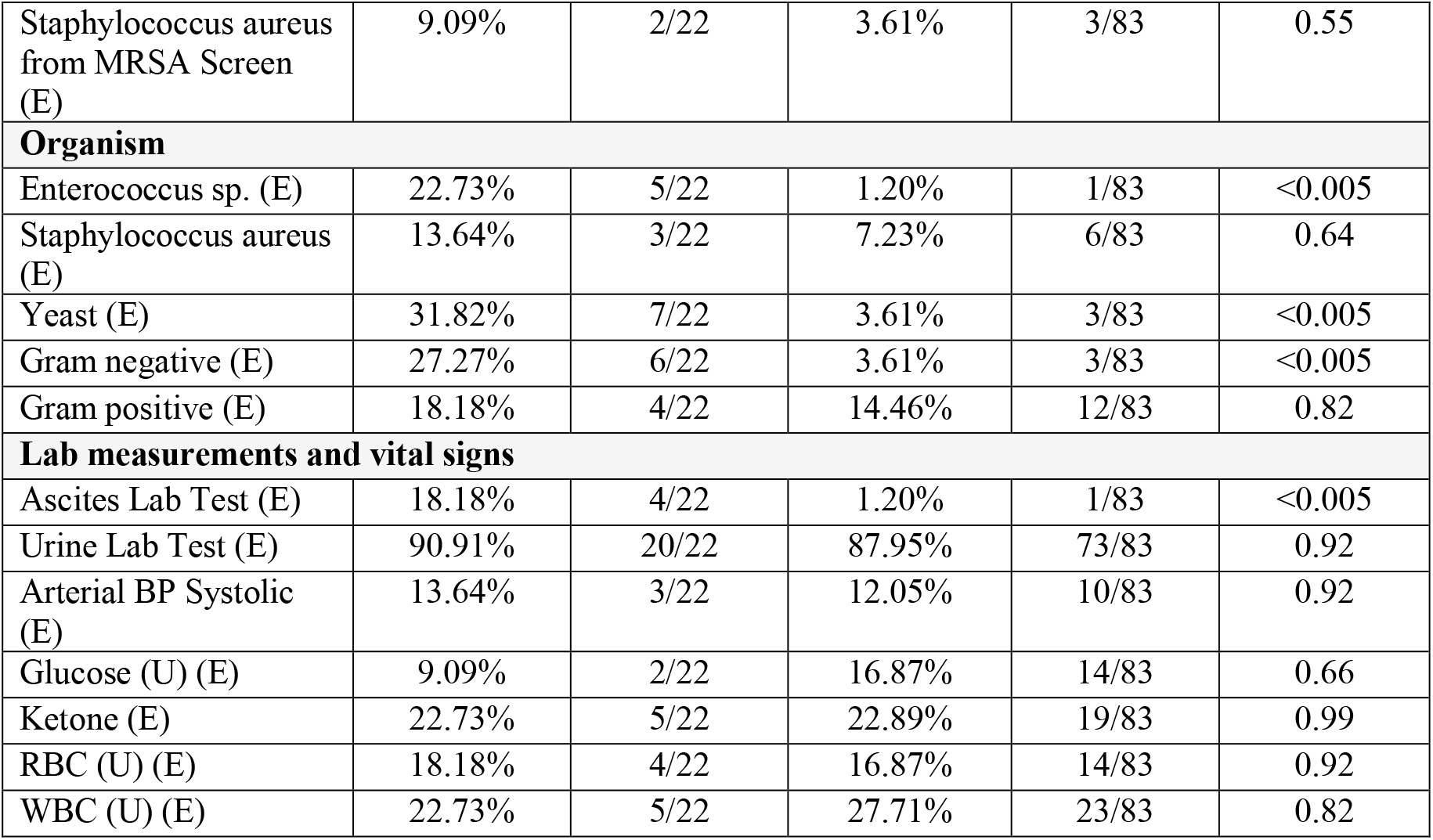
Statistics of the categorical features. Shown are, for each feature, the percentage, the fraction of patients with the feature in each class, and an FDR-corrected p-value of a Chi-square test between the two classes. R – Resistant culture, S – Sensitive culture, E – Existence feature, i.e., whether the patient had that specific measurement or culture, U-Urine.

**Supplementary Table 4.** Results of methods for removing inhuman values and outliers. The values removed by z-score and IQR methods were calculated relative to the number of values after the inhuman range filtration. The number of values removed in each step is written in parentheses. U – Urine, B – Blood.

|  | N | Inhuman removal |  |  | Z score | IQR |
| --- | --- | --- | --- | --- | --- | --- |
|  |  | Min | Max | % Values Removed | % Values Removed | % Values Removed |
| Alanine Aminotransferase (IU/L) | 358 | 0 | 20000 | 0.0% (0) | 3.1% (11) | 16.8% (60) |
| Albumin (g/dL) | 221 | 1 | 10 | 0.0% (0) | 1.8% (4) | 0.0% (0) |
| Alkaline Phosphatase (IU/L) | 346 | 10 | 200 | 8.4% (29) | 3.2% (10) | 0.0% (0) |
| Amylase (IU/L) | 167 | 0 | 1000 | 0.6% (1) | 4.8% (8) | 9.0% (15) |
| Anion Gap (mEq/L) | 1447 | 2 | 30 | 0.8% (11) | 3.6% (52) | 4.8% (69) |
| Arterial BP Systolic (mmHg) | 749 | 20 | 400 | 0.1% (1) | 4.3% (32) | 1.1% (8) |
| Arterial pH (pH) | 2080 | 6.6 | 7.8 | 0.0% (1) | 4.3% (89) | 3.5% (72) |
| Aspartate Aminotransferase (IU/L) | 359 | 0 | 20000 | 0.3% (1) | 1.7% (6) | 19.3% (69) |
| BUN (mg/dL) | 1484 | 0.1 | 200 | 0.1% (1) | 6.3% (93) | 7.8% (115) |
| Base Excess (mEq/L) | 1765 | -15 | 15 | 1.0% (18) | 4.1% (71) | 2.4% (42) |
| Basophils (%) | 165 | 0 | 10 | 0.0% (0) | 8.5% (14) | 10.9% (18) |
| Bicarbonate (mEq/L) | 1490 | 0 | 100 | 0.0% (0) | 3.7% (55) | 3.0% (45) |
| CO2 (mEq/L) | 2808 | 0 | 100 | 0.0% (1) | 3.5% (97) | 1.9% (52) |
| CaO2 (ml/dl) | 325 |  |  | 0.0% (0) | 3.4% (11) | 1.8% (6) |
| Calcium (mg/dL) | 1899 | 0 | 20 | 0.0% (0) | 3.4% (65) | 2.0% (38) |
| Chloride (mEq/L) | 1700 | 80 | 130 | 0.0% (0) | 3.5% (60) | 1.5% (25) |
| Creatine Kinase (CK) (IU/L) | 278 | 10 | 100000 | 0.0% (0) | 4.3% (12) | 13.3% (37) |
| Creatine Kinase, MB Isoenzyme (ng/mL) | 171 | 0 | 100 | 2.3% (4) | 7.2% (12) | 9.6% (16) |
| Creatinine (mg/dL) | 1491 | 0.1 | 15 | 0.0% (0) | 4.6% (69) | 9.3% (139) |
| Eosinophils (%) | 165 | 0 | 20 | 0.0% (0) | 4.2% (7) | 9.1% (15) |
| Glucose (mg/dL) | 4834 | 20 | 1000 | 0.0% (1) | 3.8% (183) | 3.9% (190) |
| Glucose (U) (mg/dL) | 20 | 100 | 1000 | 0.0% (0) | 0.0% (0) | 0.0% (0) |
| Heart Rate (BPM) | 16691 | 15 | 300 | 0.0% (5) | 3.3% (554) | 0.9% (148) |
| Hematocrit (%) | 2649 | 20 | 60 | 0.8% (22) | 3.3% (88) | 1.3% (35) |
| Hemoglobin (g/dL) | 2178 | 2 | 25 | 0.0% (0) | 3.3% (72) | 1.0% (21) |
| INR | 1005 | 0.5 | 10 | 0.4% (4) | 3.7% (37) | 10.2% (102) |
| Ionized Calcium (mmol/L) | 1083 | 0.1 | 25 | 0.0% (0) | 1.3% (14) | 4.6% (50) |
| Ketone (mg/dL) | 37 | 10 | 150 | 0.0% (0) | 13.5% (5) | 13.5% (5) |
| Lactate (mmol/L) | 904 | 0.2 | 15 | 0.4% (4) | 5.1% (46) | 6.8% (61) |
| Lactate Dehydrogenase (IU/L) | 192 | 10 | 10000 | 1.0% (2) | 3.2% (6) | 9.5% (18) |
| Lipase (IU/L) | 146 | 5 | 1000 | 4.1% (6) | 4.3% (6) | 15.0% (21) |
| Lymphocytes, Ascites (%) | 11 | 0.1 | 100 | 9.1% (1) | 0.0% (0) | 0.0% (0) |
| Lymphocytes (B) (%) | 165 | 0.1 | 100 | 1.8% (3) | 2.5% (4) | 4.3% (7) |
| MCH (pg) | 1155 | 20 | 40 | 0.0% (0) | 5.1% (59) | 4.1% (47) |
| MCHC (%) | 1156 | 25 | 45 | 0.0% (0) | 2.6% (30) | 0.7% (8) |
| MCV (fL) | 1155 | 60 | 120 | 0.0% (0) | 3.2% (37) | 2.2% (25) |
| Magnesium (mg/dL) | 1436 | 0.1 | 5 | 0.1% (1) | 4.3% (61) | 4.3% (61) |
| Monocytes, Ascites (%) | 11 | 0.1 | 22 | 63.6% (7) | 0.0% (0) | 0.0% (0) |
| Monocytes (B) (%) | 165 | 0.1 | 22 | 4.2% (7) | 5.7% (9) | 8.9% (14) |
| NBP Diastolic (mmHg) | 18886 | 20 | 400 | 0.3% (53) | 3.5% (653) | 1.9% (359) |
| NBP Mean (mmHg) | 18876 | 20 | 400 | 0.1% (27) | 3.4% (636) | 1.7% (327) |
| NBP Systolic (mmHg) | 16866 | 20 | 400 | 0.2% (37) | 3.2% (544) | 0.9% (145) |
| Neutrophils (%) | 165 | 0.2 | 100 | 1.8% (3) | 4.3% (7) | 4.3% (7) |
| Oxygen Saturation (%) | 17221 | 0 | 300 | 0.0% (0) | 1.6% (269) | 3.3% (562) |
| PEEP Set (cmH2O) | 2378 | 0 | 20 | 0.0% (0) | 5.3% (125) | 1.5% (35) |
| PT (sec) | 1004 | 5 | 35 | 2.3% (23) | 6.5% (64) | 9.3% (91) |
| PTT (sec) | 1077 | 5 | 200 | 0.0% (0) | 4.9% (53) | 7.2% (78) |
| Phosphorous (mEq/L) | 1911 | 1.5 | 7 | 5.2% (99) | 4.4% (79) | 3.6% (65) |
| Platelets (K/uL) | 1393 | 0.1 | 1000 | 0.0% (0) | 4.5% (62) | 2.6% (36) |
| Polys (%) | 11 |  |  | 0.0% (0) | 9.1% (1) | 0.0% (0) |
| Potassium (mEq/L) | 2033 | 2.5 | 9 | 0.1% (3) | 3.7% (75) | 2.8% (57) |
| Protein (mg/dL) | 70 |  |  | 0.0% (0) | 10.0% (7) | 11.4% (8) |
| RBC, Ascites (#/CU MM) | 11 |  |  | 0.0% (0) | 9.1% (1) | 18.2% (2) |
| RBC (U) (#/hpf) | 55 | 1 | 10 | 60.0% (33) | 4.5% (1) | 9.1% (2) |
| RDW (%) | 1155 | 0.1 | 50 | 0.0% (0) | 2.5% (29) | 2.3% (26) |
| Red Blood Cells (m/uL) | 1847 | 1 | 10 | 0.0% (0) | 3.4% (62) | 1.5% (28) |
| Respiratory Rate (BPM) | 18277 | 5 | 40 | 1.2% (226) | 3.4% (618) | 1.2% (216) |
| Sodium (mEq/L) | 1941 | 100 | 200 | 0.0% (0) | 4.9% (96) | 4.9% (96) |
| Specific Gravity | 187 | 0.8 | 1.2 | 0.0% (0) | 3.2% (6) | 3.7% (7) |
| Temperature C (Deg. C) | 10489 | 20 | 43 | 0.0% (5) | 4.1% (434) | 2.0% (205) |
| Total Bilirubin (mg/dL) | 412 | 0 | 40 | 0.0% (0) | 7.8% (32) | 12.4% (51) |
| Troponin T (ng/mL) | 148 |  |  | 0.0% (0) | 6.8% (10) | 15.5% (23) |
| WBC, Ascites (#/CU MM) | 11 |  |  | 0.0% (0) | 0.0% (0) | 0.0% (0) |
| WBC (U) (#/hpf) | 70 | 0.2 | 100 | 44.3% (31) | 2.6% (1) | 15.4% (6) |
| White Blood Cells (K/uL) | 1321 | 0.2 | 100 | 1.8% (24) | 3.2% (42) | 3.4% (44) |
| pCO2 (mmHg) | 1933 | 0 | 150 | 0.1% (2) | 3.5% (67) | 4.4% (85) |
| pH (U) (pH) | 196 | 3 | 7 | 7.1% (14) | 0.0% (0) | 0.0% (0) |
| pO2 (mmHg) | 1784 | 0 | 1000 | 0.0% (0) | 5.0% (90) | 7.3% (130) |

**Supplementary Table 5.** Mapping of procedures and their MIMIC-III “itemid” (in parenthesis) to the four procedure classes used by our model.

| Procedure Category | Procedure Name |
| --- | --- |
| Arterial Line | Arterial Line (225752) |
| Catheter | Dialysis Catheter (224270), ICP Catheter (226124), PA Catheter (224560), Unplanned Line/Catheter Removal (Patient Initiated) (225821), Unplanned Line/Catheter Removal (Non-Patient initiated) (225476), Pheresis Catheter (225203), Presep Catheter (224273) |
| Tube | Intubation (224385), Extubation (227194), Unplanned Extubation (patient-initiated) (225468), Unplanned Extubation (non-patient initiated) (225477), Chest Tube Placed (225433), Chest Tube Removed (227712) |
| Ventilation | Invasive Ventilation (225792), Non-invasive Ventilation (225794) |

**Supplementary Table 6.** The final model’s hyperparameters evaluated. Names and values of the Random Forest hyperparameters chosen to optimize are shown as named by Sklearn.

| Hyperparameter Name | Values Checked | Value Chosen |
| --- | --- | --- |
| n_estimators | 100, 200, 500 | 100 |
| max_features | "sqrt", "log2", None | "sqrt" |
| oob_score | False, True | True |
| max_depth | None, 4, 8 | 8 |
| class_weight | See Supplementary Table 7 | None |
| criterion | "gini", "entropy" | "gini" |
| min_samples_split | 2, 12 | 2 |
| min_samples_leaf | 1, 2, 5 | 1 |
| min_impurity_decrease | 0, 1e-5 | 1e-5 |

**Supplementary Table 7.** List of the different class weights we tried using in our models.

|  | Type of class weight |
| --- | --- |
| 1 | None (Equal weights to both classes) |
| 2 | Balanced: $\frac{totalSamples}{2 \cdot classSamples}$ |
| 3 | $(w1 = x, \quad w2 = 1 - x) \text{ for } x \text{ in } (0.75, 0.95)$ |
| 4 | $\frac{1}{classSamples}$ |
| 5 | $\frac{1}{\sqrt{classSamples}}$ |
| 6 | $\frac{1 - \beta}{1 - \beta^{classSamples}} \text{ for } \beta \text{ in } (0.9, 0.99)$ |

**Supplementary Table 8.** Average performance of the three distance methods for KNN imputation as evaluated on five splits of 5-fold cross-validation of the training set. The best performance in each category is marked in bold.

|  | Default Distance | Mean Distance Penalty | Normalization by Count of Shared Features |
| --- | --- | --- | --- |
| Mean Running Time (Seconds) | 2.292 | 2.676 | **1.189** |
| Mean AUPR (%) | 58.7 | 59.9 | **60.6** |
| Mean AUROC (%) | **82.8** | 81.3 | **82.8** |

## Supplementary text

### Feature Engineering

For lab measurements and vital signs, if the patient had more than *n* values of the feature in the relevant time frame, we fitted a linear regression model of the feature’s values over the relevant timeframe and extracted the *R*^2^ and coefficient of that model. After evaluating the impact of *n* = 3,4 and 5 on the model performance, *n* = 5 was chosen. We also created two features to compare between the values in the time-frames of *d* + 2 and *d* days: (1) the ratio between the linear regression coefficient fitted on each time-frame, (2) the difference between the median values in each time-frame. Furthermore, in order to observe the effect of the antibiotic administered to the patient, we added as a feature the ratio of the patient’s first measurement after and before the blood culture is taken.

In addition, for each lab measurement and vital sign, we created a feature of the last value recorded before PT. Since this value can be recorded at any point after the patient was admitted to the hospital, we also created a feature called “12 hours before PT”. This feature is extracted from the time window of 11-13 hours before PT. Values that were not measured between 11 to 13 hours before PT were imputed (see ‘Data Imputation Section’).

Moreover, for each lab measurements and vital sign extracted, we collected the number of measurements recorded divided by the patient’s length of stay until PT. Overall, 24 features were created for each lab test and vital sign. Moreover, an additional feature based on the temperature measurements was created, referring to the proportion of fever measurements (≥ 37.5?) out of all temperature measurements a patient had.

Furthermore, we created a binary feature for each lab measurements and vital signs feature that represents whether the patient’s feature value was imputed (i.e., masking features). In addition, for continuous lab measurements and vital signs, we added a feature that estimates how much a patient’s continuous measurements are irregular by applying Isolation Forest^52^, a method for anomaly detection.

For culture features we created features for different culture outcomes. The properties included Gram-negative, Gram-positive, the detected organism, specimen site (e.g., blood culture, sputum sample), a pair of specimen site and organism, a pair of antibiotic tested and resistance result (R for resistance, or S for sensitive), and combination of an organism, antibiotic tested and resistance result (e.g. existence of *Vancomycin*-R-*Enterococcus sp*.). We also added features for the total number of previous cultures and for total number of cultures that were found to be resistant to any antibiotic. It is important to note that our data contains the time the culture was taken, but not the time the results were retrieved. Therefore, to avoid leakage, we used only information on cultures that were taken three days or more before PT, as it takes up to three days to receive culture results. Since these features were sparse, we filtered out features with < 4% of existing values, keeping 25 features out of the original 197. The list of these 25 features appears below.

### List of previous culture features used by our model

‘AMPICILLIN - R (E)’, ‘Culture from MRSA Screen (E)’, ‘Culture from Sputum (E)’, ‘Culture from Swab (E)’, ‘Culture from Urine (E)’, ‘Enterococcus sp. - AMPICILLIN - R (E)’, ‘Enterococcus sp. - PENICILLIN - R (E)’, ‘Enterococcus sp. - VANCOMYCIN - R (E)’, ‘Enterococcus sp. from Swab (E)’, ‘Enterococcus sp. (E)’, ‘GENTAMICIN - S (E)’, ‘LEVOFLOXACIN - R (E)’, ‘OXACILLIN - R (E)’, ‘PENICILLIN - R (E)’, ‘Staphylococcus aureus - OXACILLIN - R (E)’, ‘Staphylococcus aureus from MRSA Screen (E)’, ‘Staphylococcus aureus (E)’, ‘VANCOMYCIN - R (E)’, ‘VANCOMYCIN - S (E)’, ‘Gram Negative (E)’, ‘Gram Positive (E)’, ‘Yeast from Urine (E)’, ‘Yeast (E)’.

R – Resistant culture, S – Sensitive culture, E – existence feature, i.e., whether the patient had that specific result.

### Data Imputation

For data imputation, we used KNN and compared three different distance methods. The first metric we used was Sklearn’s weighted distance metric^53,54^ defined as

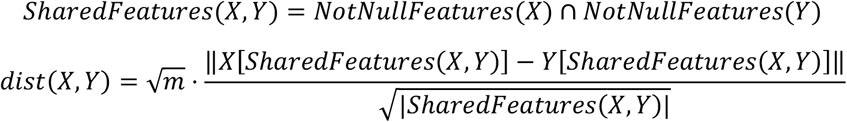

The imputed value of the feature *l* in a patient with feature vector *X* is 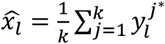, where 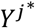 is the feature vector of its j^th^ nearest neighbor.

In the second distance metric, “Mean Distance Penalty”, we added a penalty to the distance calculation for each feature that is missing in either vector. Define *penalty*_*f*_ as the mean square distance calculated between non missing values of feature *f*. For efficiency, we used in the computation 10% of the non-missing values of the feature, sampled from evenly-spaced quantiles of the feature. Then define

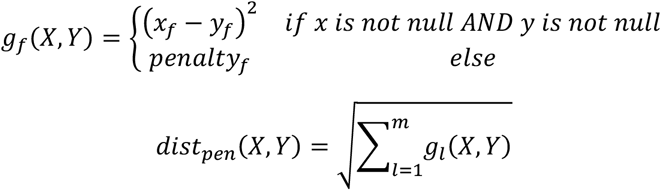

In the third method, named “Normalization by Count of Shared Features”, we normalized the default distance method by the number of not-null feature values shared by the two vectors instead of normalizing by the squared root of this number, as follows:

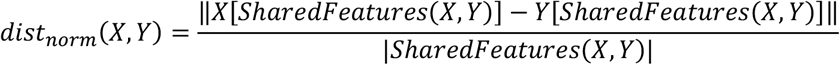

This gives more weight to the number of not-null values than the default method.

We then evaluated the effect on the model performance and the running time of each distance method (**Supplementary Table 8**). Based on these results, we chose the third distance function.

## References

1. Hornischer, K. & Häußler, S. Diagnostics and Resistance Profiling of Bacterial Pathogens. in (eds. Stadler, M. & Dersch, P.) 89–102 (Springer International Publishing, 2016).

2. Bell, B. G., Schellevis, F., Stobberingh, E., Goossens, H. & Pringle, M. A systematic review and meta-analysis of the effects of antibiotic consumption on antibiotic resistance. BMC Infectious Diseases 14, 13 (2014).

3. Wall, S. Prevention of antibiotic resistance – an epidemiological scoping review to identify research categories and knowledge gaps. Global Health Action 12, 1756191 (2019).

4. Laxminarayan, R. et al. Antibiotic resistance—the need for global solutions. The Lancet Infectious Diseases 13, 1057–1098 (2013).

5. Nathan, C. & Cars, O. Antibiotic Resistance — Problems, Progress, and Prospects. New England Journal of Medicine 371, 1761–1763 (2014).

6. Aslam, B. et al. Antibiotic resistance: a rundown of a global crisis. Infect Drug Resist 11, 1645–1658 (2018).

7. Mendelson, M. Review: Role of antibiotic stewardship in extending the age of modern medicine. South African Medical Journal 105, 414–419 (2015).

8. Niederman, M. S. Appropriate use of antimicrobial agents: Challenges and strategies for improvement. Critical Care Medicine 31, 608 (2003).

9. Thomson, R. B. & McElvania, E. Blood Culture Results Reporting: How Fast Is Your Laboratory and Is Faster Better? Journal of Clinical Microbiology 56, e01313–18 (2018).

10. Livermore, D. M. & Wain, J. Revolutionising Bacteriology to Improve Treatment Outcomes and Antibiotic Stewardship. Infect Chemother 45, 1–10 (2013).

11. Kumar, A. Antimicrobial Delay and Outcome in Severe Sepsis. Critical Care Medicine 42, e802 (2014).

12. Kumar, A. et al. Duration of hypotension before initiation of effective antimicrobial therapy is the critical determinant of survival in human septic shock*. Critical Care Medicine 34, 1589 (2006).

13. Luyt, C.-E., Bréchot, N., Trouillet, J.-L. & Chastre, J. Antibiotic stewardship in the intensive care unit. Crit Care 18, 480 (2014).

14. Bassetti, M. et al. Systematic review of the impact of appropriate versus inappropriate initial antibiotic therapy on outcomes of patients with severe bacterial infections. International Journal of Antimicrobial Agents 56, 106184 (2020).

15. Raman, G., Avendano, E., Berger, S. & Menon, V. Appropriate initial antibiotic therapy in hospitalized patients with gram-negative infections: systematic review and meta-analysis. BMC Infect Dis 15, 395 (2015).

16. Vallés, J., Rello, J., Ochagavía, A., Garnacho, J. & Alcalá, M. A. Community-Acquired Bloodstream Infection in Critically Ill Adult Patients: Impact of Shock and Inappropriate Antibiotic Therapy on Survival. Chest 123, 1615–1624 (2003).

17. Rhodes, A. et al. Surviving Sepsis Campaign: International Guidelines for Management of Sepsis and Septic Shock: 2016. Intensive Care Med 43, 304–377 (2017).

18. Johnson, A. E. W. et al. MIMIC-III, a freely accessible critical care database. Scientific Data 3, 1–9 (2016).

19. Altman, N. S. An introduction to kernel and nearest-neighbor nonparametric regression. American Statistician 46, 175–185 (1992).

20. Guyon, I., Weston, J., Barnhill, S. & Vapnik, V. Gene Selection for Cancer Classification using Support Vector Machines. Machine Learning 46, 389–422 (2002).

21. Pedregosa, F. et al. Scikit-learn: Machine Learning in Python. MACHINE LEARNING IN PYTHON.

22. Lundberg, S. M. & Lee, S. I. A unified approach to interpreting model predictions. Advances in Neural Information Processing Systems 2017-Decem, 4766–4775 (2017).

23. He, H., Bai, Y., Garcia, E. A. & Li, S. ADASYN: Adaptive synthetic sampling approach for imbalanced learning. in 2008 IEEE International Joint Conference on Neural Networks (IEEE World Congress on Computational Intelligence) 1322–1328 (2008).

24. Chawla, N. V., Bowyer, K. W., Hall, L. O. & Kegelmeyer, W. P. SMOTE: Synthetic Minority Over-sampling Technique. Journal of Artificial Intelligence Research 16, 321–357 (2002).

25. Han, H., Wang, W.-Y. & Mao, B.-H. Borderline-SMOTE: A New Over-Sampling Method in Imbalanced Data Sets Learning BT - Advances in Intelligent Computing. in (eds. Huang, D.-S., Zhang, X.-P. & Huang, G.-B.) 878–887 (Springer Berlin Heidelberg, 2005).

26. Breiman, L. Random Forests. Machinelearning202.Pbworks.Com 1–35 (1999).

27. Freund, Y. & Schapire, R. E. A Decision-Theoretic Generalization of On-Line Learning and an Application to Boosting. Journal of Computer and System Sciences 55, 119–139 (1997).

28. Cox, D. R. The Regression Analysis of Binary Sequences. Journal of the Royal Statistical Society. Series B (Methodological) 20, 215–242 (1958).

29. Cortes, C. & Vapnik, V. Support-vector networks. Machine Learning 20, 273–297 (1995).

30. Ke, G. et al. LightGBM: A Highly Efficient Gradient Boosting Decision Tree. in Advances in Neural Information Processing Systems vol. 30 (Curran Associates, Inc., 2017).

31. Friedman, J. H. Stochastic gradient boosting. Computational Statistics & Data Analysis 38, 367–378 (2002).

32. Chen, T. & Guestrin, C. XGBoost: A Scalable Tree Boosting System. in Proceedings of the 22nd ACM SIGKDD International Conference on Knowledge Discovery and Data Mining 785–794 (Association for Computing Machinery, 2016). doi:10.1145/2939672.2939785.

33. Bilavsky, E., Yarden-Bilavsky, H., Ashkenazi, S. & Amir, J. C-reactive protein as a marker of serious bacterial infections in hospitalized febrile infants. Acta Paediatrica 98, 1776–1780 (2009).

34. Rasmussen, N. H. & Rasmussen, L. N. Predictive Value of White Blood Cell Count and Differential Cell Count to Bacterial Infections in Children. Acta Paediatrica 71, 775–778 (1982).

35. Brown, L., Shaw, T. & Wittlake, W. A. Does leucocytosis identify bacterial infections in febrile neonates presenting to the emergency department? Emergency Medicine Journal 22, 256–259 (2005).

36. Vincent, J.-L. et al. International Study of the Prevalence and Outcomes of Infection in Intensive Care Units. JAMA 302, 2323–2329 (2009).

37. Tabah, A. et al. Characteristics and determinants of outcome of hospital-acquired bloodstream infections in intensive care units: the EUROBACT International Cohort Study. Intensive Care Med 38, 1930–1945 (2012).

38. Roimi, M. et al. Early diagnosis of bloodstream infections in the intensive care unit using machine-learning algorithms. Intensive Care Med 46, 454–462 (2020).

39. Zoabi, Y. et al. Predicting bloodstream infection outcome using machine learning. Sci Rep 11, 20101 (2021).

40. Goodman, K. E. et al. A Clinical Decision Tree to Predict Whether a Bacteremic Patient Is Infected With an Extended-Spectrum β-Lactamase–Producing Organism. Clin Infect Dis 63, 896–903 (2016).

41. Oonsivilai, M. et al. Using machine learning to guide targeted and locally-tailored empiric antibiotic prescribing in a children’s hospital in Cambodia. Wellcome Open Res 3, 131 (2018).

42. Yelin, I. et al. Personal clinical history predicts antibiotic resistance of urinary tract infections. Nat Med 25, 1143–1152 (2019).

43. Kanjilal, S. et al. A decision algorithm to promote outpatient antimicrobial stewardship for uncomplicated urinary tract infection. Science Translational Medicine 12, eaay5067 (2020).

44. MacFadden, D. R. et al. Utility of prior cultures in predicting antibiotic resistance of bloodstream infections due to Gram-negative pathogens: a multicentre observational cohort study. Clinical Microbiology and Infection 24, 493–499 (2018).

45. Chatterjee, A. et al. Quantifying drivers of antibiotic resistance in humans: a systematic review. The Lancet Infectious Diseases 18, e368–e378 (2018).

46. Vazquez-Guillamet, M. C., Vazquez, R., Micek, S. T. & Kollef, M. H. Predicting Resistance to Piperacillin-Tazobactam, Cefepime and Meropenem in Septic Patients With Bloodstream Infection Due to Gram-Negative Bacteria. Clinical Infectious Diseases 65, 1607–1614 (2017).

47. Lewin-Epstein, O., Baruch, S., Hadany, L., Stein, G. Y. & Obolski, U. Predicting Antibiotic Resistance in Hospitalized Patients by Applying Machine Learning to Electronic Medical Records. Clinical Infectious Diseases 72, e848–e855 (2021).

48. Hernàndez-Carnerero, À. et al. Dimensionality reduction and ensemble of LSTMs for antimicrobial resistance prediction. Artificial Intelligence in Medicine 138, 102508 (2023).

49. Previsdomini, M., Gini, M., Cerutti, B., Dolina, M. & Perren, A. Predictors of positive blood cultures in critically ill patients: a retrospective evaluation. Croat Med J 53, 30–39 (2012).

50. Agarwal, M. & Larson, E. L. Risk of drug resistance in repeat gram-negative infections among patients with multiple hospitalizations. J Crit Care 43, 260–264 (2018).

51. Johnson, A., Pollard, T. & Mark, R. MIMIC-III Clinical Database. (2015).

52. Liu, F. T., Ting, K. M. & Zhou, Z. H. Isolation forest. Proceedings - IEEE International Conference on Data Mining, ICDM 413–422 (2008) doi:10.1109/ICDM.2008.17.

53. Pedregosa, F. et al. Scikit-learn: Machine Learning in Python. MACHINE LEARNING IN PYTHON.

54. Dixon, J. K. Pattern Recognition with Partly Missing Data. IEEE Transactions on Systems, Man, and Cybernetics 9, 617–621 (1979).

